# Baseline expression of immune gene modules in blood is associated with primary response to anti-TNF therapy in Crohn’s disease patients

**DOI:** 10.1101/2023.04.19.23288234

**Authors:** Benjamin Y. H. Bai, Mark Reppell, Nizar Smaoui, Jeffery F. Waring, Valerie Pivorunas, Heath Guay, Simeng Lin, Neil Chanchlani, Claire Bewshea, James R. Goodhand, UK Inflammatory Bowel Disease Pharmacogenetics Study Group, Nicholas A. Kennedy, Tariq Ahmad, Carl A. Anderson

## Abstract

**Background and Aims:** Anti-TNF therapy is widely used for treatment of inflammatory bowel disease (IBD), yet many patients are primary non-responders (PNR), failing to respond to induction therapy. We aimed to identify blood gene expression differences between primary responders (PR) and primary non-responders to anti-TNF monoclonal antibodies (infliximab and adalimumab); and to predict response status from blood gene expression and clinical data.

**Methods:** The Personalised Anti-TNF Therapy in Crohn’s Disease (PANTS) study is a UK-wide prospective observational cohort study of anti-TNF therapy outcome in anti-TNF naive Crohn’s disease (CD) patients (ClinicalTrials.gov identifier: NCT03088449). Blood gene expression in 324 unique patients was measured by RNA-seq at baseline (week 0), and at weeks 14, 30, and 54 after treatment initiation (total sample size = 814).

**Results:** After adjusting for clinical covariates and estimated blood cell composition, baseline expression of MHC, antigen presentation, myeloid cell enriched receptor, and other innate immune gene modules was significantly higher in anti-TNF responders. Expression changes from baseline to week 14 were generally of consistent direction across patients but with greater magnitude in responders, with the exception of interferon-related genes that were upregulated uniquely in non-responders. Expression differences between responders and non-responders observed at week 14 were maintained at week 30 and week 54. Prediction of response status from baseline clinical data, cell composition, and module expression was poor.

**Conclusions:** Baseline gene module expression was associated with primary response to anti-TNF therapy in PANTS patients. However, these expression differences were insufficient for clinically useful prediction of response.

## Introduction

Crohn’s disease (CD) is a chronic immune-mediated inflammatory disease (IMID) of the gastrointestinal tract. Along with ulcerative colitis (UC), it is one of the two main forms of inflammatory bowel disease (IBD). The development of anti-TNF biologic therapies has revolutionised patient care for CD and a number of other IMIDs over the last two decades. Two major anti-TNF drugs, infliximab and adalimumab, are IgG1 monoclonal antibodies that bind both soluble and transmembrane TNF, inhibiting their interactions with TNF receptors.^1, 2^ Two main mechanisms of action have been proposed: induction of CD4^+^ T cell apoptosis in the gut mucosa by inhibiting the TNF-TNFR2 interaction; and binding of the antibody tail (Fc region) of the drug to Fc receptors on monocytes, inducing their differentiation into wound-healing M2 macrophages.^3, 4^

Unfortunately, anti-TNF therapy is not always effective at treating IBD. Various types of treatment failure can occur: primary non-response (PNR) within the induction period (the first 12–14 weeks for infliximab and adalimumab), secondary loss of response (LOR) during maintenance therapy after an initial response, failure to achieve remission after the treatment course, or adverse events that lead to treatment discontinuation.^5^ For IBD patients, the incidence of PNR is 10–40%, and the incidence of secondary LOR among initial responders is 24–46% in the first year of treatment.^6–8^ The ability to predict PNR and LOR could help guide changes in treatment regimens, such as dose intensification or switching to a drug class with a different mechanism of action.^2, 6^ Reliable baseline prediction would be especially valuable, allowing stratification of patients to effective therapies from treatment initiation, minimising healthcare costs and patient burden.

Clinical variables reported to be associated with anti-TNF response include age, disease duration, body mass index (BMI), smoking, C-reactive protein (CRP) levels, faecal calprotectin levels, serum drug concentrations, and anti-drug antibody concentrations. These associations have mostly been reported in small retrospective cohorts and have rarely been independently validated.^7, 9–13^ In the Personalised Anti-TNF Therapy in Crohn’s Disease (PANTS) study, the largest study of infliximab and adalimumab response in CD patients to date (enrolment n = 1610), baseline obesity, smoking, and greater disease activity were associated with low serum drug concentration after induction. Low drug concentration was in turn associated with PNR and non-remission, suggesting immunogenicity may be mediating treatment failure by increasing drug clearance rate.^8^

Multiple studies have attempted to define transcriptomic predictors for anti-TNF response.^12–20^ Reported associations include TREM1 expression, identified as a marker of anti-TNF response in different studies with inconsistent directions of effect. Gaujoux et al.^16^ found TREM1 expression was lower in gut biopsies from infliximab responders than in non-responders (total cohort size n = 72), but higher in responders in a separate cohort measuring baseline whole blood expression (n = 22). By contrast, Verstockt et al.^18^ reported lower TREM1 expression in responders to infliximab and adalimumab in both baseline gut biopsies (n = 44) and baseline whole blood (n = 54). Proposed reasons for the discrepancy include false positives due to small sample sizes, differences in patient ethnicity, and differing definitions of response.^12, 21^ In general, small sample sizes, and variation among studies in analysis methods, anti-TNF drug, response definition, tissues sampled, and disease subtype make a consensus hard to establish. Few markers for anti-TNF response of any type, clinical or transcriptomic, have been validated in independent studies, and none have yet been translated to routine clinical practice.^13^

To identify novel transcriptomic associations with primary response to anti-TNF therapy, we generated longitudinal RNA-seq data from peripheral blood samples taken from a subset of the PANTS cohort (182 PR, 142 PNR) during the first year of follow-up. Differential gene expression (DGE) between primary responders and non-responders was performed at baseline (week 0), post-induction (week 14), and during maintenance (week 30 and week 54). We detected differences in gene module expression that may reflect differences in disease characteristics or severity that influence risk of primary non-response. As this is one of the largest datasets currently available for assessing transcriptomic associations with anti-TNF response in IBD, we also examined the significance of previously reported transcriptomic markers from the literature. Finally, we evaluated the utility of measuring module expression for prediction of primary response status.

## Materials and Methods

### Study design

PANTS is a UK-wide, multicentre, prospective observational cohort study reporting the treatment failure rates of the anti-TNF drugs infliximab (originator, Remicade [Merck Sharp & Dohme, Hertfordshire, UK] and biosimilar, CT-P13 [Celltrion, Incheon, South Korea]), and adalimumab (Humira [AbbVie, Cambridge, MA]) in 1610 anti-TNF-naive patients with active luminal CD. The study design has been described in detail previously.^8, 55^ In brief, patients were recruited at the time of first anti-TNF exposure between February 2013 and June 2016, and evaluated for 12 months or until drug withdrawal. Eligible patients were aged ≥ 6 years with evidence of active luminal Crohn’s disease involving the colon and/or small intestine. Four major study visits were scheduled at first dose (baseline, week 0), post-induction (week 14), week 30, and week 54. Additional visits were scheduled at treatment failure or exit. At baseline, clinical and demographic data were recorded, including sex, ethnicity, BMI, smoking status, age at diagnosis, disease duration, Montreal classification, prior medical and drug history, and previous Crohn’s disease-related surgeries. At every visit, disease activity score, weight, current therapy, and adverse events were recorded.

### RNA-seq sample selection

A subset of PANTS patients was selected for RNA-seq, with the inclusion criteria: age ≥ 16 years; and baseline CRP ≥ 4 mg/L and/or baseline calprotectin > 100 µg/g. The target sample size was 200 patients on infliximab and 200 patients on adalimumab, with an even split between PR and PNR within each drug group. PR and PNR were defined based on Kennedy et al.^8^ patient outcome criteria:

- Primary non-response (assessed at week 14): exit before week 14 because of treatment failure (including resectional inflammatory bowel disease surgery) or corticosteroid use at week 14 (new prescriptions or if previous dose had not been stopped). Patients whose CRP did not decrease to 3 mg/L or less or by 50% or more from baseline (week 0), and whose HBI score did not decrease to 4 points or less or by 3 points or more from baseline, were also classified as having a primary non-response.
- Primary response (assessed at week 14): decrease in CRP to 3 mg/L or less or by 50% or more from baseline (week 0) and a decrease in HBI to 4 points or less or by 3 points or more from baseline.
- Remission (assessed at weeks 14, 30, 54; implies primary response): CRP of 3 mg/L or less and HBI score of 4 points or less, no ongoing steroid therapy, and no exit due to treatment failure.

Steroid use was defined as any systemic therapy, either oral or intravenous (including use of steroids for other conditions), but excluding single pre-infusion dosing with hydrocortisone.

PNR were required to exhibit primary non-response at week 14 and non-remission at week 54. PR were required to exhibit primary response or remission at week 14, and be in remission at week 54 (or week 30 if week 54 status was unknown). Furthermore, within infliximab-treated patients, PNR and non-PNR were matched based on baseline immunomodulator use, baseline steroid use, age at first dose, baseline albumin, sex, and weight at study entry.

### Whole blood RNA-seq

Whole blood was collected in RNA Tempus tubes (Applied Biosystems) and stored at −80°C until extraction (QIAsymphony PAXgene Blood RNA Kit, Qiagen). RNA was quantified using the QuBit BR RNA (ThermoFisher), and RNA integrity was assessed with the 4200 TapeStation (Agilent). RNA-seq libraries were prepared using the Kapa mRNA HyperPrep Kit, with depletion of ribosomal RNA (rRNA) and globin mRNA using the QIAseq FastSelect RNA Removal Kit, and adapter ligation with IDT xGEN Dual Index UMI adapters. A total of 1 141 samples from 396 patients were sequenced. Raw sequencing data was demultiplexed with Picard^22^ and aligned to the reference genome (GRCh38) using STAR (v2.6.1d)^23^. Reads were deduplicated using UMI-tools^24^ and quantified against the Ensembl 96 gene annotation with featureCounts (v1.6.4).^25^

Outlier samples were excluded, defined as > 2 standard deviations from the mean based on percentage of aligned reads in coding regions reported by Picard, percentage of unique reads and number of unique reads. Samples with sex mismatch against documented sex were removed. As gene expression measured from bulk tissue is heavily dependent on cell composition,^26^ cell proportions of six common cell types in whole blood (CD4^+^ T cells, CD8^+^ T cells, B cells, NK cells, monocytes, and granulocytes) were estimated using the Houseman method^27^ from paired DNA methylation data.^28^ Samples missing clinical data and/or cell proportion estimates were removed. A total of 814 samples remained after filtering. To accommodate variability in sampling day, samples were mapped to timepoints based on Kennedy et al.^8^ windows around major visits: week 0 (week −4–0), week 14 (week 10–20), week 30 (week 22–38), and week 54 (week 42–66). Samples taken at additional visits (LOR or exit) falling within one of the windows were mapped to that timepoint, unless the patient also had a major visit sample inside that window. The mapping of samples to timepoints is shown in Fig. S1a. The number of samples per patient ranged from one to four, with a median of three (Fig. S1b).

Counts were normalised for library size using edgeR (v3.28.1).^29^ Globin genes and short non-coding RNAs were removed, and genes with low expression were filtered, requiring genes to have at least 1.25 counts per million in > 10% of samples and non-zero expression in > 90% of samples. Expression data from 15 511 genes remained after filtering. Finally, log2 expression values were generated using variancePartition/voom.^30, 31^

### Statistical analyses

A full description of the statistical analyses can be found in the Supplementary Methods. In brief, DGE analyses were performed in R (v3.6.2),^32^ with the significance threshold set at false discovery rate (FDR) < 0.05. Variance components analysis was used to identify influential variables for inclusion in DGE models (Fig. S2). Cell proportion estimates were found to explain large fractions of expression variance, and adjusting for cell proportions reduced the number of significant associations but improved consistency between drug subgroups, with fewer highly significant modules showing significant drug-by-response interaction effects compared to the unadjusted results (Fig. S3). As this study was not designed to compare between drug subgroups, we focused on models adjusted for cell composition, where the improved consistency allows us to pool expression data from both subgroups for greater statistical power. For all DGE models, cell proportions, sequencing batch, age of onset (the patient’s age at disease diagnosis), disease duration, body mass index (BMI), anti-TNF drug type, prior surgery, and smoking were included as fixed effects; and patient was included as a random effect.

Per-gene linear mixed-effects models fit using DREAM (variancePartition v1.16.1)^33^ were used to detect pairwise DGE between study groups. Additionally, natural cubic splines (splines::ns)^32^ were fit to explore non-linear expression trajectories over all four timepoints, modelling expression as a function of study day in each group (first drug dose = study day 0). Different expression trajectories were detected by testing for differences in spline parameters between groups. Significant genes from the spline analysis were hierarchically clustered by their mean expression in PR and PNR at each timepoint, and the gap statistic^34^ was used to define clusters of genes with distinct trajectories. The spline analyses were only performed with drug subgroups pooled, as relatively small sample sizes at week 30 and week 54 precluded stratification by drug.

Rank-based gene set enrichment analyses (tmod::tmodCERNOtest, v0.46.2),^35^ using blood transcriptomic modules (BTMs) were performed to identify coordinately up or downregulated gene sets. These modules are sets of genes that are coexpressed in whole blood, derived by Li et al.^36^ (module names prefixed with “LI”) and Chaussabel et al.^37^ (prefixed “DC”) from publicly available expression datasets. Gene set overrepresentation analyses were run for BTMs (tmod::tmodHGtest) and other publicly-available gene sets (gprofiler2::gost, v0.2.0).^38^

Single-sample gene set enrichment scores (ssGSEA, https://github.com/broadinstitute/ssGSEA2.0/) were computed as a summary measure of module expression in a sample, both at baseline and at week 14. Predictive models using clinical variables, cell proportions, and module expression scores (baseline or week 14) to predict response status were constructed using caret (v6.0-86).^39^ Multiple predictive algorithms were evaluated: penalised and regularised logistic regression methods, parallel random forest, eXtreme Gradient Boosting, support vector machines with a radial basis, k-nearest neighbours, naive Bayes, and Gaussian process models. Bootstrapping (50 replicates) with the AUC metric was used to tune models, evaluate internal performance, and perform model selection. Pairwise tests for the difference in AUCs were performed with pROC.^40^

### Ethical statement

The South West Research Ethics committee approved the study (Research Ethics Committee reference: 12/SW/0323) in January 2013. Patients were included after providing informed, written consent. The study is registered with ClinicalTrials.gov identifier NCT03088449, and the protocol is available at https://www.ibdresearch.co.uk/pants/.

## Results

### Baseline expression modules are associated with post-induction primary non-response

After RNA-seq quantification and quality control, expression data was available for 15 511 genes and 814 samples. These samples were from 324 patients, whose characteristics are shown in Table 1. No single gene had a significant association between week 0 expression and response in the infliximab subgroup (86 PR, 59 PNR). Expression of NK cell (LI.M7.2) and T cell (LI.M7.1, LI.M7.0) gene modules (BTMs)^36^ was significantly lower in responders. Exclusively in the adalimumab subgroup (66 PR, 57 PNR), PDIA5 (log2 fold change (FC) = - 0.3512, FDR = 0.006777), KCNN3 (log2 FC = −0.8798, FDR = 0.006777), and IGKV1-9 (log2 FC = −1.223, FDR = 0.04518) had significantly lower expression in responders (Fig. 1a). This was accompanied by lower expression of plasma cell/immunoglobulin (LI.M156.0, LI.M156.1) and cell cycle (LI.M4.0, LI.M4.1) modules (Fig. 1b). This heterogeneity between drug subgroups was robust to model form (Fig. S4) and differences in sample size between subgroups (Fig. S5). A pooled analysis was performed to identify modules consistently differentially expressed in both drug subgroups (152 PR, 116 PNR). MHC-TLR7-TLR8 cluster (LI.M146), antigen presentation (LI.M71, LI.M95.0), and myeloid cell enriched receptor and transporter (LI.M4.3) modules were found to be upregulated at baseline in responders. We also examined previously reported baseline markers in gut mucosal biopsies and blood from the literature,^14, 15, 18, 41^ and did not find them to be significant in this study (Fig. 1a).

**Figure 1.**
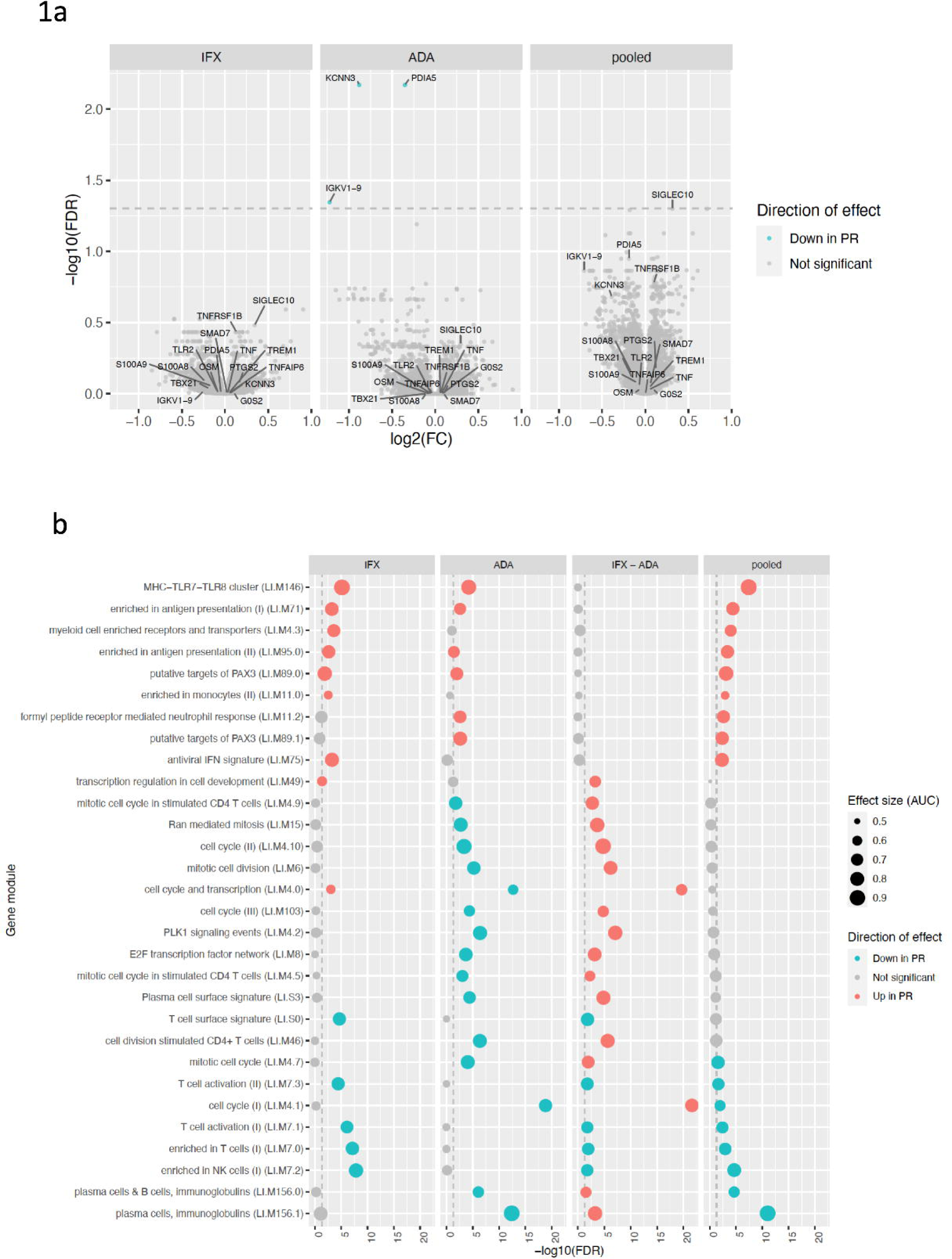
Baseline expression associated with primary response, adjusted for cell composition. a. Volcano plots of DGE between responders (PR) and non-responders (PNR) at week 0; for infliximab (IFX), adalimumab (ADA), or with drug subgroups pooled. Annotated genes show significant associations from this study, and previously reported associations from the literature in both blood and gut biopsies. Dashed line shows significance threshold at FDR = 0.05. b. Top gene modules differentially expressed between PR and PNR at week 0. Columns correspond to results for infliximab (IFX), adalimumab (ADA), difference between IFX and ADA (IFX − ADA i.e. drug-by-response interaction), and pooled drug analyses. The top 30 modules ranked by minimum FDR in any column are shown. Dashed lines show significance thresholds at FDR = 0.05.

**Table 1.** Baseline patient characteristics for the PANTS RNA-seq subcohort. Characteristics are stratified by drug subgroup. Values shown are count and percentage for categorical variables, and mean and standard deviation for continuous variables. Nominal p-values are reported for the comparison between drug subgroups.

|  | adalimumab (ADA) | infliximab (IFX) | drugs pooled | *p*-value |
| --- | --- | --- | --- | --- |
| **Sex** |  |  |  | 0.317 |
| (Col %) |  |  |  | Fisher exact |
| FEMALE | 78 (48.4%) | 89 (54.6%) | 167 (51.5%) |  |
| MALE | 83 (51.6%) | 74 (45.4%) | 157 (48.5%) |  |
| **Age of onset (years)** |  |  |  | 0.774 |
| Mean (SD) | 33.3 (15.4) | 32.8 (15.3) | 33.1 (15.3) | Wilcoxon rank-sum |
| Missing | 0 | 0 | 0 |  |
| **Disease duration (years)** |  |  |  | 0.546 |
| Mean (SD) | 6.1 (8.1) | 5.9 (7.7) | 6.0 (7.9) | Wilcoxon rank-sum |
| Missing | 0 | 0 | 0 |  |
| **Smoking status** |  |  |  | 0.263 |
| (Col %) |  |  |  | Fisher exact |
| Current | 28 (17.4%) | 36 (22.1%) | 64 (19.8%) |  |
| Ex | 55 (34.2%) | 43 (26.4%) | 98 (30.2%) |  |
| Never | 78 (48.4%) | 84 (51.5%) | 162 (50.0%) |  |
| **Crohn's-related surgery** |  |  |  | 0.549 |
| (Col %) |  |  |  | Fisher exact |
| NO | 114 (70.8%) | 110 (67.5%) | 224 (69.1%) |  |
| YES | 47 (29.2%) | 53 (32.5%) | 100 (30.9%) |  |
| **On immunomodulator ever** |  |  |  | 0.543 |
| (Col %) |  |  |  | Fisher exact |
| NO | 23 (14.3%) | 28 (17.2%) | 51 (15.7%) |  |
| YES | 138 (85.7%) | 135 (82.8%) | 273 (84.3%) |  |
| **On immunomodulator at baseline** |  |  |  | 0.912 |
| (Col %) |  |  |  | Fisher exact |
| NO | 79 (49.1%) | 81 (49.7%) | 160 (49.4%) |  |
| YES | 82 (50.9%) | 82 (50.3%) | 164 (50.6%) |  |
| **On corticosteroids at baseline** |  |  |  | 0.011 |
| (Col %) |  |  |  | Fisher exact |
| NO | 113 (70.2%) | 92 (56.4%) | 205 (63.3%) |  |
| YES | 48 (29.8%) | 71 (43.6%) | 119 (36.7%) |  |
| **Baseline BMI** |  |  |  | 0.237 |
| Mean (SD) | 25.2 (6.2) | 24.3 (5.5) | 24.8 (5.9) | Wilcoxon rank-sum |
| Missing | 0 | 0 | 0 |  |
| **Primary response status** |  |  |  | 0.263 |
| (Col %) |  |  |  | Fisher exact |
| Primary non-response | 76 (47.2%) | 66 (40.5%) | 142 (43.8%) |  |
| Primary response | 85 (52.8%) | 97 (59.5%) | 182 (56.2%) |  |
| **CD8<sup>+</sup> T cell (%)** |  |  |  | 0.380 |
| Mean (SD) | 2.8 (4.2) | 2.8 (5.2) | 2.8 (4.7) | Wilcoxon rank-sum |
| Missing | 38 | 18 | 56 |  |
| **CD4<sup>+</sup> T cell (%)** |  |  |  | 0.752 |
| Mean (SD) | 9.2 (6.3) | 9.2 (6.8) | 9.2 (6.5) | Wilcoxon rank-sum |
| Missing | 38 | 18 | 56 |  |
| **B cell (%)** |  |  |  | 0.094 |
| Mean (SD) | 1.9 (2.0) | 1.5 (1.9) | 1.7 (1.9) | Wilcoxon rank-sum |
| Missing | 38 | 18 | 56 |  |
| **Monocyte (%)** |  |  |  | 0.497 |
| Mean (SD) | 8.9 (3.5) | 9.2 (3.7) | 9.0 (3.6) | Wilcoxon rank-sum |
| Missing | 38 | 18 | 56 |  |
| **NK cell (%)** |  |  |  | 0.683 |
| Mean (SD) | 1.9 (3.2) | 1.9 (3.8) | 1.9 (3.5) | Wilcoxon rank-sum |
| Missing | 38 | 18 | 56 |  |
| **Granulocyte (%)** |  |  |  | 0.911 |
| Mean (SD) | 74.3 (9.7) | 74.3 (10.8) | 74.3 (10.3) | Wilcoxon rank-sum |
| Missing | 38 | 18 | 56 |  |

### Expression changes from baseline to post-induction are largely amplified in primary responders

To characterise the changes in gene expression induced by anti-TNF therapy, we compared expression at baseline to expression post-induction, and also estimated the difference between expression changes in responders and non-responders (the timepoint-by-response interaction). As expression changes from week 0 to week 14 were relatively consistent between patients on infliximab and adalimumab after adjusting for cell composition (Fig. S6), we pooled drug subgroups for these models. We found that 5 572 and 626 genes were differentially expressed in responders and non-responders respectively, with 179 genes having a significant timepoint-by-response interaction. Of the genes differentially expressed between week 14 and week 0 in both responders and non-responders, and with a significant timepoint-by-response interaction, nearly all (31/32 genes) had an expression change that was amplified in responders (Fig. 2a). For example, CD177, a neutrophil marker upregulated during inflammation, was downregulated at week 14 in responders to a much greater extent (log2 FC = −2.225, FDR = 4.104 × 10^−17^) than in non-responders (log2 FC = −0.8981, FDR = 0.004598; interaction FDR = 0.008247). Modules differentially expressed between week 0 to week 14 included upregulation of B cell (LI.M47.0), plasma cell (LI.M156.0), and T cell activation (LI.M7.1) modules; and downregulation of immune activation (LI.M37.0), monocyte (LI.M11.0), neutrophil (LI.M37.1), and TLR and inflammatory signalling (LI.M16) modules (Fig. 2b). Amplification of expression changes in responders was also observed at the module level, with nearly all module expression changes aligned in the same direction in responders and non-responders, but with larger effects in responders.

**Figure 2.**
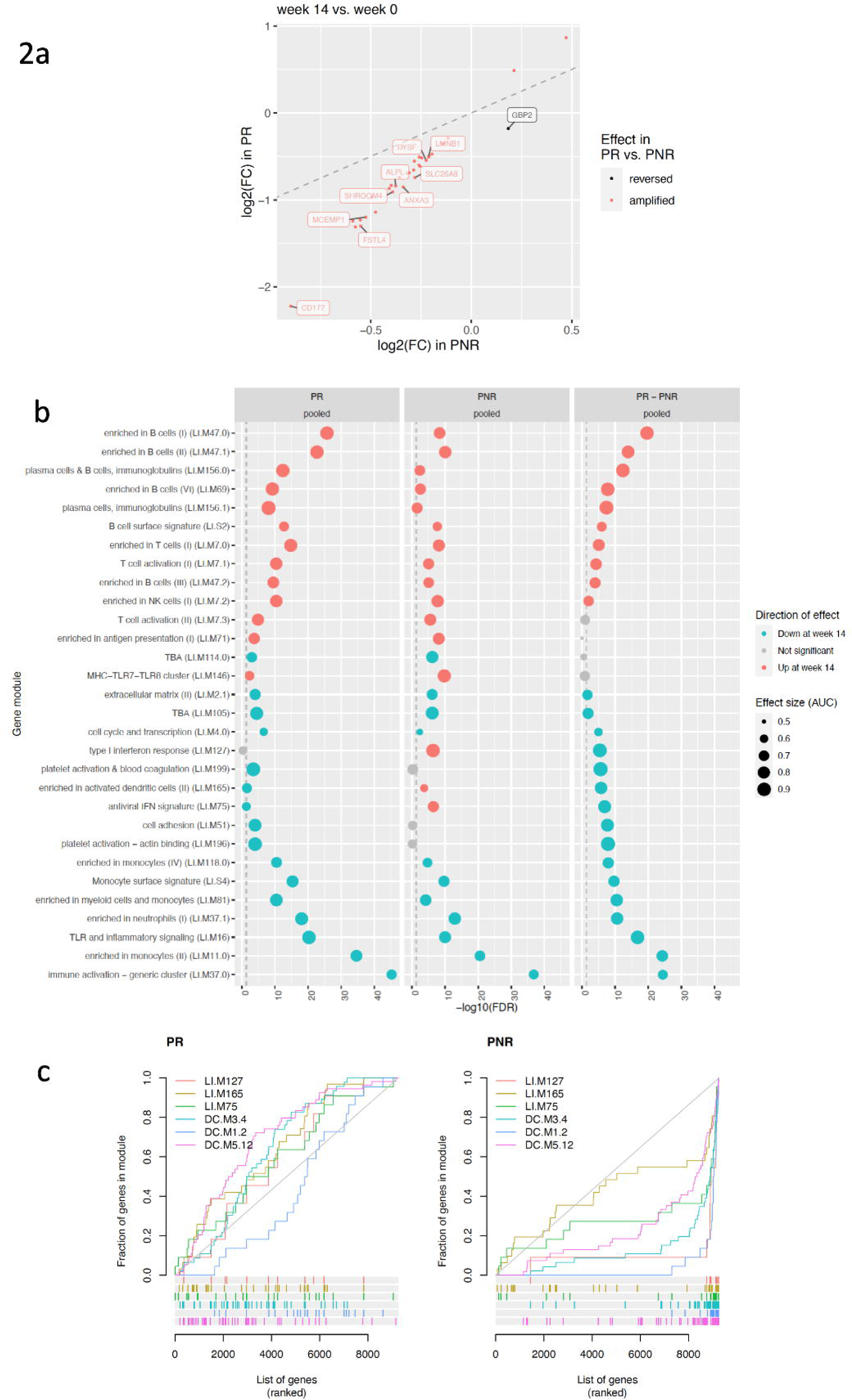
Expression changes from baseline to post-induction are amplified in responders. a. Expression log2 fold changes from week 0 to week 14 in primary responders (PR) and non-responders (PNR), for genes that differentially expressed from week 0 to week 14 in both responders and non-responders, with a significantly different effect size between responders and non-responders (top 10 labelled). The identity line is shown by the dashed line. b. Top modules differentially expressed between week 14 and week 0, adjusted for cell composition. Columns show effects in primary responders (PR), non-responders (PNR), and the primary responder minus non-responder difference. The top 30 modules ranked by minimum FDR in any column are shown. Vertical dashed line shows significance threshold at FDR = 0.05. c. Barcode plots showing interferon modules (LI/DC) specifically upregulated from week 0 to week 14 in primary non-responders (PNR), but not in primary responders (PR). Genes were ranked in ascending order by week 14 versus week 0 DGE z-statistic. Curves show the cumulative fraction of genes in each module. Effect sizes are represented by the area under the curves (AUCs). Diagonal line shows the null of randomly-distributed ranks.

In contrast, GBP2 (a member of a family of guanylate-binding proteins induced by interferons^42^) was downregulated from week 0 to week 14 in responders (log2 FC = −0.1783, FDR = 0.004878), but upregulated in non-responders (log2 FC = 0.1849, FDR = 0.04502; interaction FDR = 0.005977). At the module level, upregulation of type I interferon response (LI.M127), activated dendritic cell (LI.M165), and antiviral IFN signature (LI.M75) modules was observed in non-responders but not in responders (Fig. 2b). Further gene set enrichment analyses using modules from Chaussabel et al.^37^ also revealed annotated interferon gene modules to be significantly upregulated at week 14 in non-responders: DC.M3.4, containing STAT2, GBP5, and PARP14 (FDR = 3.447 × 10^−21^); as well two modules containing IFIT3 and GBP2, DC.M1.2 (FDR = 9.492 × 10^−16^) and DC.M5.12 (FDR = 1.355 × 10^−13^). None of these modules were differentially expressed from week 0 to week 14 in responders (Fig. 2c), suggesting upregulation of interferon pathways at week 14 occurs uniquely in primary non-responders to anti-TNF.

### Sustained expression differences between responders and non-responders during maintenance

As PANTS was an observational study, it was possible to retain patients who continued with anti-TNF therapy even after meeting the study definition of PNR at week 14, enabling us to sample the blood transcriptome at week 30 and week 54 during the maintenance period. Leveraging all 814 samples over the four study timepoints, we tested for general differences in expression trajectory over time, detecting 210 differentially expressed genes between responders and non-responders after adjustment for cell composition. To visualise the expression of these genes and identify common patterns of expression change during anti-TNF therapy, significant genes were hierarchically clustered by their expression. Six clusters were identified (Fig. 3a), each with distinct expression trajectories for responders and non-responders (Fig. 3b). Cluster 1 largely comprised genes previously found to have a significant difference in expression change from week 0 to week 14 between responders and non-responders (97/132 genes). The most significant gene was KREMEN1 (FDR = 4.287 × 10^−4^), part of an inflammatory apoptotic pathway in gut epithelium.^43^ Cluster 1 genes were enriched in modules associated with myeloid cells and monocytes (LI.M81, hypergeometric test, FDR = 2.115 × 10^−6^), platelet activation (LI.M196, 1.348 × 10^−5^), immune activation (LI.M37.0, 1.436 × 10^−4^), and TLR and inflammatory signalling (LI.M16, FDR = 2.365 × 10^−3^) (Fig. 3c). Expression trajectories showed cluster 1 genes were more downregulated from baseline in responders than non-responders, likely representing a lower inflammatory state in responders by week 14 that is also maintained at week 30 and week 54. An opposing trend was observed in cluster 5, which contained genes enriched for B cell development/activation (LI.M58, FDR = 0.01653) that were more upregulated from baseline in responders than non-responders.

**Figure 3.**
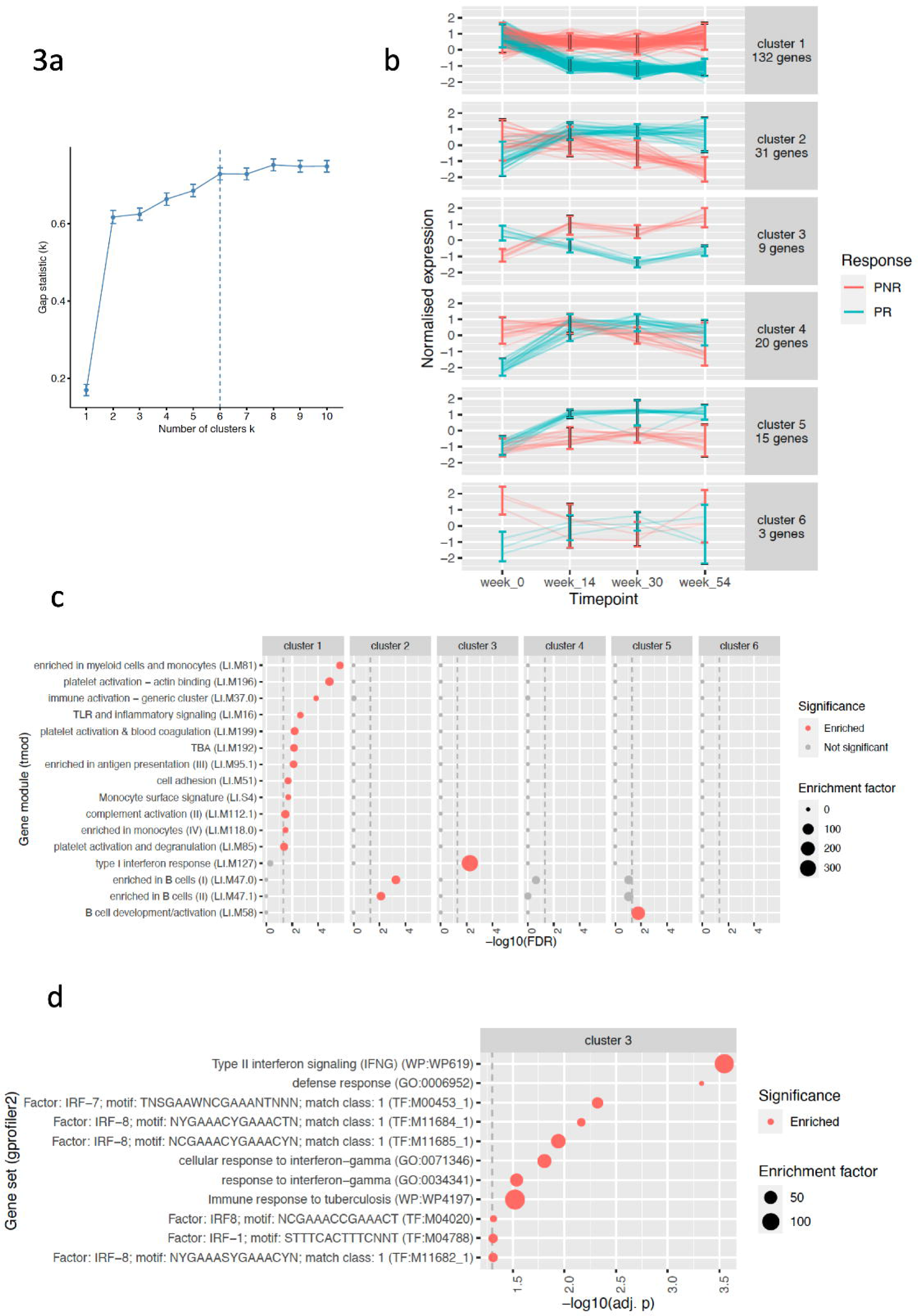
Sustained expression differences between responders and non-responders during maintenance. a. Gap statistic versus cluster number k from hierarchical clustering of genes with significant expression differences between PR and PNR over all timepoints. Error bars derived from 500 bootstraps. The optimal cluster number was selected to be k = 6 by the factoextra::fviz_nbclust firstSEmax criteria (https://rpkgs.datanovia.com/factoextra/index.html). b. Normalised expression over study timepoints for genes in each cluster. 95% confidence intervals for expression are shown for each group at each timepoint. c. Gene modules enriched in each cluster from gene set overrepresentation analyses using tmod::tmodHGtest. Modules significantly enriched in any cluster are shown. Vertical dashed line shows significance threshold at FDR = 0.05. d. Gene sets enriched in cluster 3 from gene set overrepresentation analyses using gprofiler2::gost. Vertical dashed line shows significance threshold at adjusted p-value = 0.05 (gost g:SCS multiple testing correction method).

Cluster 3 was uniquely enriched for type I interferon response genes (LI.M127, FDR = 0.005681, Fig. 3c). Subsequent enrichment analyses using publicly-available gene sets^38^ revealed enrichments for type II interferon signalling (WP:WP619, adj. p = 0.0002826), and for genes containing putative transcription factor (TF) binding motifs for interferon regulatory factors IRF7 (TF:M00453_1, adj. p = 0.004768) and IRF8 (TF:M11684_1, adj. p = 0.006853; TF:M11685_1, adj. p = 0.01136) (Fig. 3d). The genes in cluster 3 showed opposing directions of expression change from week 0 to week 14 in responders versus non-responders, generating expression differences at week 14 that were sustained at week 30 and week 54. Of the nine genes in the cluster, eight genes (STAT1, BATF2, GBP1, GBP5, IRF1, TAP1, APOL1, APOL2) had a significant interaction between week 0 to week 14 expression change and response status in the per-gene differential expression analyses. IRF1 and STAT1 are key transcription factors for interferon signalling.^44, 45^ Unlike the majority of genes that followed trajectories of greater expression change in responders, genes in interferon response pathways were uniquely upregulated in non-responders.

### Prediction of primary non-response from gene expression and clinical variables

Given there were multiple significant module-level associations with primary response status, we evaluated the ability to predict response from module expression, as well as cell proportions and clinical variables. Single-sample gene set enrichment scores (ssGSEA) were computed to summarise module expression per sample. Using baseline module scores, the median resampling AUCs over all combinations of algorithms and predictor sets ranged between 0.5541 and 0.6686 (Fig. S7). The best-performing model was regularised logistic regression (regLogistic; hyperparameters: cost = 0.25, loss = L1, epsilon = 0.01) using clinical variables, cell proportions, and module scores as predictors, giving a median resampling AUC of 0.6686, a median sensitivity of 0.5392, and a median specificity of 0.6852, where non-response was the positive class, with a prevalence of 43% (116/268). Including cell proportions and module scores improved predictive performance compared to the model using only clinical variables (bootstrap p = 0.02629), but the increase in AUC was only 2.5% (Fig. 4a). This suggests that clinical variables provided the greatest contributions to baseline prediction, especially those variables with high importance in all three models (high absolute t-statistics): smoking history, BMI, and baseline steroid usage (Fig. 4b).

**Figure 4.**
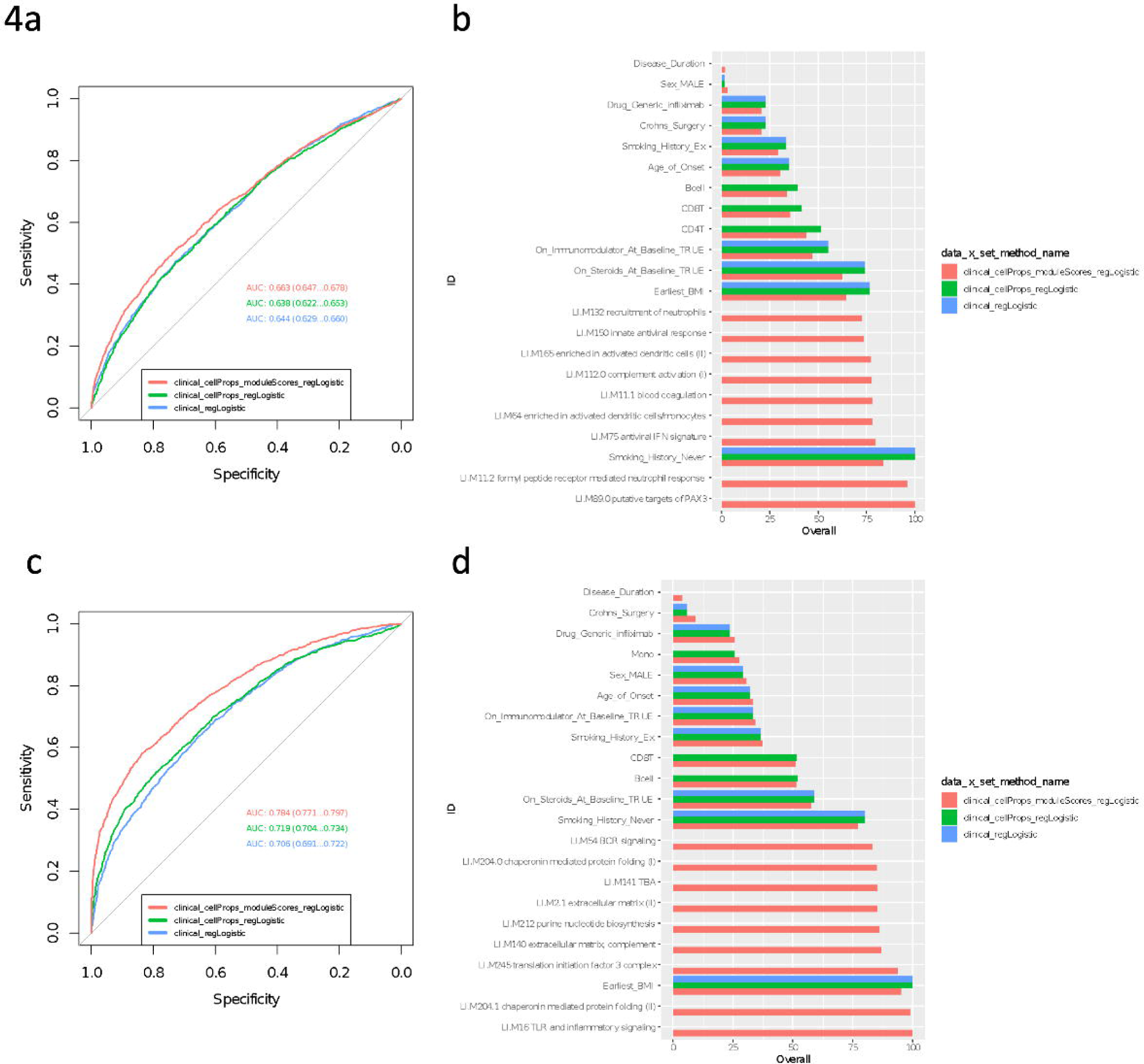
Prediction of response status from clinical variables, cell proportions, and expression data. ROC curves for the regLogistic method trained on each predictor dataset are shown at baseline (a) and week 14 (c). ROC curves were plotted after merging all 50 resamples. Primary non-response was used as the positive class. DeLong 95% confidence intervals for the AUC are shown. The ten most important variables from models trained on each predictor dataset are shown for baseline (b) and week 14 (d) models. The overall variable importance score is computed from the absolute value of the t-statistic for each predictor from the final tuned models. Missing bars denote variable that were not in the predictor dataset for that model.

Performance for predicting primary non-response could be improved by using week 14 cell proportions and module scores (median resampling AUC range 0.6216-0.7957, Fig. S8). The prevalence of non-response at week 14 was 42% (104/246). The best performing predictor was again regularised logistic regression incorporating clinical variables, cell proportions, and module scores as predictors (median resampling AUC = 0.7957, sensitivity = 0.6248, specificity = 0.7961). Here the addition of module scores to the predictor dataset had a larger benefit on predictive performance, with a 6.5% increase in AUC comparing the full model to the model including only clinical variables and cell proportions (bootstrap p = 2.306 × 10^−10^), with a (Fig. 4c). The week 14 modules with the highest variable importance included TLR and inflammatory signalling (LI.M16), chaperonin mediated protein folding (LI.M204.0, LI.M204.1) and translation initiation factor 3 complex (LI.M245) modules (Fig. 4d). Greater predictive performance at week 14 likely reflects the larger clinical and expression differences observed between responders and non-responders after the induction period.

## Discussion

We found substantial differences in whole blood gene expression between anti-TNF primary responders and non-responders in the PANTS cohort. At baseline, three single-gene associations detected in the adalimumab subgroup implicated similar cell types; IGKV1-9 encodes the immunoglobulin light chain variable region that forms part of antibodies produced by plasma cells, KCNN3 is annotated to a plasma cell surface signature module from Li et al.^36^ (LI.S3), and the expression of both KCNN3 and PDIA5 is high in plasma cells (www.proteinatlas.org/humanproteome/immune+cell, v21.1) and positively correlated with plasmablast frequencies in blood.^46^ These genes were downregulated in responders, as was the expression of plasma cell and immunoglobulin modules. In keeping with our observations, Martin et al.^17^ identified plasma cells as part of a correlated module of cell populations, where lower module expression in gut biopsies was associated with better response to anti-TNFs. Baseline plasma cell abundances in gut biopsies have also been reported to be lower in responders, albeit in relatively small cohorts of infliximab patients.^16^ Our findings lend credence that associations driven by immune cells observed in gut biopsies may also be observable in blood, a more accessible tissue.

Previously reported single-gene baseline markers in gut biopsies and blood were non-significant in this study. For example, TREM1^16, 18^ was not significantly differentially expressed between responders and non-responders in blood samples from PANTS patients. Our observation is consistent with two recent trials of comparable sample size, SERENE-CD and SERENE-UC, where baseline blood TREM1 expression was not predictive of response in either CD or UC patients.^47^ A variety of factors could explain failures to replicate reported markers from study to study. Many existing studies pool heterogeneous cohorts of patients taking different anti-TNF drugs due to the scarcity of large datasets; even between arms of the PANTS study, we observe within-study differences in expression. Additional between-study variation arises from differences in clinical setting, tissues sampled (e.g. blood versus gut biopsies), and definition of primary response (e.g. endoscopic versus clinical parameters). Any two studies are unlikely to have adjusted for the same combinations of covariates in modelling, including covariates such as cell composition that are influential for bulk expression data. Finally, small sample sizes have considerable sampling error. Despite only finding a small number of single-gene associations in PANTS, we detected multiple novel module-level associations, such as baseline upregulation of MHC-mediated antigen presentation modules in responders. We highly recommend the use of set-based methods that draw on changes in multiple genes, as they are not only better powered, but may be more reproducible compared to single-gene association tests.

Leveraging the longitudinal design of PANTS, we characterised the changes in blood gene expression post-induction. Reduced expression of immune activation modules in responders at week 14 is consistent with successful drug inhibition of TNF-mediated inflammation. Decreased inflammation correlates with reduced neutrophil activation and reduced monocyte recruitment,^48^ and apoptosis of monocytes induced by anti-TNF in CD patients has been previously described.^43^ Certain B cell subsets are reduced in the blood of IBD patients compared to controls,^49^ so upregulation of B cell modules at week 14 may also represent a shift towards health. Similar expression changes were observed in responders and non-responders, but with greater magnitude in responders, potentially supporting the hypothesis that response is a continuum. Gaujoux et al.^16^ found changes in cell proportions in response to anti-TNF treatment were amplified in responders; here we demonstrate a similar trend at the transcriptional level in PANTS. Post-induction expression differences between responders and non-responders were sustained at week 30 and week 54 during the anti-TNF maintenance period. Kennedy et al.^8^ found that “continuing standard dosing regimens after primary non-response was rarely helpful” for inducing remission by week 54. This phenomenon may also be reflected in the blood transcriptome, although non-responders in the PANTS RNA-seq data were selected to exclude patients in remission by week 54, so trajectories for non-responders at week 14 that eventually achieved remission could not be observed. Thus, the observed difference in trajectories between responders and non-responders is likely exaggerated.

Unlike the majority of baseline versus post-induction associations, expression changes in genes and modules in the interferon pathway were uniquely upregulated in PANTS non-responders. Previous studies in IBD are conflicting, with Samie et al.^50^ reporting elevated expression of interferon pathway genes in colonic biopsies from non-responders compared to responders, with no significant change pre-versus post-treatment (n ≈ 40); and Mavragani et al.^20^ reporting a post-treatment reduction in blood interferon expression only in non-responders (n = 30). In studies of rheumatoid arthritis (RA), another IMID with licensed anti-TNF therapies, increases in type I interferon-regulated gene expression in blood after infliximab treatment were associated with poor clinical response (discovery n = 15, validation n = 18).^51^ A systematic review of our study with other studies reporting similar associations between interferon pathway genes and anti-TNF response would not only help resolve the direction of effect, if any, but provide an opportunity to consider the shared biology of anti-TNF response in IBD, RA, and other immune-mediated diseases.

An important limitation of our analyses is that PANTS was not designed to compare between drug subgroups. Differences between patients on different anti-TNF drugs can arise from patient and physician preference, influenced by cost, disease severity, location, and comorbidities. Unsurprisingly, many associations with response had significantly different effect sizes in infliximab and adalimumab patient subgroups. We found that adjusting DGE models for estimated proportions of major cell types as a proxy for these uncontrolled factors alleviated heterogeneity between subgroups. However, the adjustment is unlikely to work well for rare cell types, thus the associations we report may reflect differences in cell proportions rather than per-cell expression. Given the myriad of other factors that could drive the remaining heterogeneity, we strongly caution against interpreting associations with different effects in the PANTS infliximab and adalimumab subgroups as drug-driven differences with biological significance, and recommend that future transcriptomic studies consider influential factors such as cell composition.

We were unable to build clinically useful predictive models of response incorporating expression data. Using only baseline clinical variables, Kennedy et al.^8^ used logistic regression with stepwise variable selection based on Akaike information criterion (AIC) to predict response in the full PANTS cohort, achieving AUCs of 0.53 (95% CI 0.46–0.59) for infliximab patients and 0.54 (0.46–0.62) for adalimumab patients. Whilst our best-performing baseline model represents an improvement, expression data only contributed a small amount of predictive power on top of clinical variables and cell composition. Unsurprisingly, models had greater predictive power when provided with week 14 expression and cell composition data, and adding expression data also provided a comparatively large increase in AUC. This suggests that when expression differences between responders and non-responders are sufficiently large, transcriptomic markers do provide unique information, and are not simply proxies for clinical variables or coarse estimates of blood cell composition. A potential route to more effective prediction is to consider whether expression differences arising early in the induction period can discriminate between responders and non-responders. For example, Mesko et al.^52^ found that week 2 blood gene expression was predictive of infliximab response in CD (discovery n = 20, validation n = 20) and RA (discovery n = 19, validation n = 15) patients. More recently, Mishra et al.^53^ trained random forest models using blood DNA methylation and gene expression measured in IBD patients receiving infliximab (n = 37). They did not find consistent baseline-only predictive signatures, but a model combining baseline with week 2 measurements predicted response in the Mesko et al. cohort with 85% accuracy (95% CI: 62–97%). As we observed in PANTS, expression differences between responders and non-responders were far greater by week 14 than at baseline. Post-induction associations were also more consistent between drug subgroups, as baseline differences are diluted by the large transcriptomic perturbation from taking an anti-TNF. Expression changes in the innate immune system are observable within hours of treatment initiation,^53^ and robust prediction of non-response within that timeframe may be more valuable than a less reliable prediction at baseline.

In conclusion, we observed significant differences in gene module expression between responders and non-responders to anti-TNF therapy in the whole blood of PANTS CD patients at baseline and post-treatment timepoints. Interferon-induced genes were uniquely upregulated post-induction in non-responders, going against the general trend of amplified transcriptomic change in responders versus non-responders. However, there is not yet a consensus on the direction of these effects, and we were unable to robustly predict response with our current sample size. To obtain accurate prediction, especially from baseline data, leveraging large upcoming datasets with paired drug response phenotypes and transcriptomic data such as the 1000IBD project will be essential.^54^

## Author Contributions

SL, NC, JRG, NAK, TA, and CAA participated in the conception and design of the study. CB was the project manager and coordinated recruitment. BYHB, MR, NS, JFW, VP, HG, SL, NC, CB, JRG, NAK, TA, and CAA were involved in the acquisition, analysis, or interpretation of data. The data analysis was performed by BYHB, MR, SL, and NAK. Drafting of the manuscript was conducted by BYHB. All authors contributed to the revision of the manuscript for critically important intellectual content. TA obtained the funding for the study. CAA is the guarantor of the manuscript. All authors contributed to the final approval of the manuscript.

## Data Availability

Individual participant de-identified data that underlie the results reported in this article will be available immediately after publication for a period of 5 years. The data will be made available to investigators whose proposed use of the data has been approved by an independent review committee. Analyses will be restricted to the aims in the approved proposal. Proposals should be directed to Tariq Ahmad and Carl A. Anderson. To gain access data requestors will need to sign a data access agreement.

## Acknowledgements

We would like to acknowledge Jim Butler, Areej Ammar, Erin Murphy, Stephen Abel, Elizabeth Asque, and Justin Wade Davis from AbbVie Inc., Chicago, IL, USA for supporting this work. Laboratory tests were undertaken by the Exeter Blood Sciences Laboratory at the Royal Devon & Exeter (RD&E) NHS Trust (https://www.exeterlaboratory.com/). The Exeter NIHR Clinical Research Facility coordinated sample storage and management.lllThe sponsor of the study was the Royal Devon and Exeter NHS Foundation Trust. Simeng Lin is supported by alllWellcomelllGW4-CAT fellowship (222850/Z/21/Z). Neil Chanchlani acknowledges support from Crohn’s & Colitis UK. We thank the patients who participated in the PANTS study, the Inflammatory Bowel Disease Pharmacogenetic Study Group, and research nurses who collected clinical data and biological samples at each study visit. We acknowledge the study co-ordinators of the Exeter IBD Research Group, United Kingdom: Marian Parkinson and Helen Gardner-Thorpe for their ongoing administrative support to the study.

## Abbreviations

ADA: adalimumab
AIC: Akaike information criterion
BMI: body mass index
BTMs: blood transcriptomic modules
CD: Crohn’s disease
CRP: C-reactive protein
FDR: false discovery rate
IBD: inflammatory bowel disease
IFX: infliximab
IMID: immune-mediated inflammatory disease
LOR: loss of response
PANTS: Personalised Anti-TNF Therapy in Crohn’s Disease
PNR: primary non-response/non-responder
PR: primary response/responder
TNF: tumour necrosis factor
UC: ulcerative colitis
rRNA: ribosomal RNA
ssGSEA: single-sample gene set enrichment score

