## Supplementary Information for "Baseline expression of immune gene modules in blood is associated with primary response to anti-TNF therapy in Crohn’s disease patients"

#### Supplementary Methods

##### Study design

###### The PANTS study

PANTS is a UK-wide, prospective, observational cohort study of response to anti-TNF therapy in CD patients, previously described in detail by Kennedy et al.[1] and Sazonovs et al.[27] The study is registered with ClinicalTrials.gov identifier NCT03088449, and the protocol is available at https://www.ibdresearch.co.uk/pants/. Total enrolment was 1610 patients, who were at least 6 years old, had active luminal CD, and were naive to anti-TNF therapy.

The anti-TNF drugs evaluated were adalimumab (ADA), infliximab (IFX), and infliximab biosimilars. Patients received treatment according to standard dosing intervals (8 weeks for infliximab, 2 weeks for adalimumab), and were invited to attend up to ten major study visits over the course of the study, or until anti-TNF drug withdrawal. These visits were timed to allow sampling immediately prior to drug doses, and to allow the same visit structure to be used for all patients. Additional visits were scheduled in case of secondary LOR or premature study exit due to drug withdrawal, usually replacing the next scheduled major visit.

The target timings for the four major visits in the first year of follow-up in the PANTS study were week 0, week 14, week 30, and week 54. The week 0 major visit is the visit prior to the first dose of drug. Week 0 to week 14 is the induction period. After week 14, patients continued to take their drug for maintenance according to the standard dosing interval. For RNA-seq, whole blood samples at these four major visits were taken prior to the scheduled drug doses that aligned with those visits, labelled with the visit name, and preserved in Tempus Blood RNA Tubes.

#### Definition of primary response and primary non-response

The definition of primary response and non-response was based on the clinical decision tree from Kennedy et al.[1], as detailed in the main text ("RNA-seq sample selection"). Primary response and non-response could be assessed from week 12, with a final classification made by the scheduled week 14 visit. As PANTS was an observational study that continued until drug withdrawal, a patient's clinician may have decided to continue anti-TNF therapy even if a patient demonstrated primary non-response, so it was possible for primary non-responders to remain in the study past week 14.

#### Selection of samples for RNA-seq

Additional selection criteria were used to choose the subcohort of PANTS patients put forward for RNA-seq. Patients were required to be at least 16 years old, on either infliximab or adalimumab (not on an infliximab biosimilar), and to have an available baseline serum sample. Equal numbers of responders and non-responders to each drug were selected, excluding grey zone patients that only partially met the criteria for either primary non-response or response. Within the patients on infliximab, there was propensity score matching between primary non-responders and other patients based on baseline immunomodulator use, baseline steroid use, age, sex, and body mass index (BMI). Furthermore, primary non-responders were selected excluding patients known to be in remission at week 54, and primary responders were selected from patients known to be in remission by week 30 or week 54. The primary non-responders and responders in the RNA-seq subcohort thus represent phenotypic extremes of response.

It could not be guaranteed that study visits occurred on the target day specified in the protocol, thus samples were mapped to timepoints using windows around major visits defined by Kennedy et al.[1] Relative to the first dose of anti-TNF (day 0), these windows were: week 0 (week −4–0), week 14 (week 10–20), week 30 (week 22–38), and week 54 (week 42–66). Only a small minority of labelled major visit samples fell outside their respective windows, mostly for later timepoints where there was more variation around the target day. Samples taken at additional (LOR or exit) visits falling within one of the windows were mapped to that timepoint.

#### Library preparation and sequencing

Total RNA was extracted following the Qiagen QIAsymphony instrument protocol (RNA Isolation PAX RNA CR22332 ID 2915). RNA was quantified with the ThermoFisher QuBit BR RNA (Q10211), and RNA integrity assessed with the Agilent RNA ScreenTape assay (5067-5579, 5067-5577, 5067-5576) on the Agilent 4200 TapeStation. Library preparation was performed using the Kapa mRNA HyperPrep Kit, including enrichment for messenger RNA (mRNA) using magnetic oligo-dT beads, depletion of ribosomal RNA (rRNA) and globin mRNA using the QIAseq FastSelect RNA Removal Kit, and adapter ligation with IDT xGEN Dual Index UMI adapters. Libraries were sequenced on the Illumina HiSeq 4000 with 75 bp paired-end reads.

#### RNA-seq quantification and preprocessing

A total of 1141 samples from 396 patients were sequenced to a target minimum depth of 20 million total read pairs before deduplication. Sequencing data was demultiplexed with Picard.[2] Sequence quality, overrepresented sequences, adapter content, and sequence duplication rates were checked using FastQC.[3] Reads were mapped to GRCh38 using STAR (v2.6.1d)[4] and deduplicated to unique reads using UMI-tools.[5] Gene expression was quantified against the Ensembl 96 gene annotation with featureCounts (v1.6.4).[6]

Samples were filtered to remove outliers (>2 standard deviations from the mean) according to percentage of aligned reads in coding regions reported by Picard, percentage of unique reads, and number of unique reads. Samples that could not be mapped to a timepoint were removed. Samples with sex mismatch were removed. Samples from patients with grey zone primary response were removed. Samples for which there was missingness in the data matrix for clinical and cell proportion were removed. A total of 814 samples remained after filtering (Fig. S1a). The number of samples per patient ranged from one to four, with a median of three (Fig. S1b).

The Ensembl 96 gene annotation contains 58 884 genes, many of which are not expressed in whole blood. Effective library sizes were computed using the trimmed mean of M-values

(TMM) method in edgeR (v3.28.1),[7] then between-sample normalisation for library size was performed using edgeR::cpm, converting counts to counts per million (CPM). Genes with low expression were filtered, requiring >1.25 CPM in >10% of samples (1.25 CPM being approximately 10 counts at the median library size of 8 million unique mapped read pairs) and non-zero expression in >90% of samples. Globin genes and short non-coding RNAs (ncRNAs) were removed. A total of 15 511 genes remained after filtering. Finally, CPMs were converted to the log2 scale, and precision weights to account for the expression mean-variance relationship were computed for each gene and sample using variancePartition::voomWithDreamWeights.[8]

### Differential gene expression

#### Variable selection by variance components analysis

For each gene, the DGE model was a regression expressing the response variable (gene expression), as a linear function of predictor variables of interest (primary response status, drug, timepoint), and other selected predictor variables. Fig. S9 shows the correlation matrix of variables considered for selection. These included three variables from Kennedy et al.[1] associated with primary response: baseline immunomodulator use, smoking, and BMI. Also available were proportions of six common cell types in whole blood (CD4+ T cells, CD8+ T cells, B cells, NK cells, monocytes, granulocytes), estimated from whole blood Illumina MethylationEPIC methylation array data collected from the same patients and timepoints.[9] Estimates were computed using the Houseman method, which uses differentially methylated regions between immune cell types as cell type markers,[10] implemented in Minfi (minfi::estimateCellCounts).[11]

A variance components analysis was performed to quantify the proportion of expression variance explained by each variable for each gene using variancePartition,[8] as variables that do not explain much variation in the response would be unlikely to improve statistical efficiency if conditioned on in DGE models. The variance components analysis model was a linear mixed effects regression model with variables in Fig. S9 included as predictors. Additional categorical variables were included for patient and RNA-seq library preparation plate. An additional continuous variable consisting of random numbers drawn from the standard normal distribution was included as a null (random_numbers). Granulocyte proportion estimates were dropped to relieve perfect multicollinearity. Categorical variables were coded as random intercepts, and continuous variables as fixed effects—simulations from Hoffman et al.[8] showed variance proportion estimates were unbiased even when coding categorical variables with as few as two categories as random effects, as long as model parameters were estimated using maximum likelihood (ML) rather than restricted maximum likelihood (REML). It was also shown this approach avoids overestimates of variance proportions that occur if categorical variables with many levels are treated as fixed.

As downstream DGE methods required the same set of predictors for all genes, we aimed to select variables that explained substantial variance for many genes. Variables that explained the most variance on average were patient, cell proportions, and RNA-seq plate (Fig. S2). Some variables that did not explain more variance on average than the null nevertheless had high maximum values, indicating their importance for a relatively small number of genes. These included sex, library preparation protocol version (always the same within RNA-seq plates), and smoking status. However primary response status—a variable of interest—also fell into this group, so it was difficult to justify excluding all variables with lower median variance explained than the null. Consequently, all non-null variables in Fig. S2 were selected as predictors in downstream models apart from Ever_Immunomodulator (whether the patient had ever had immunomodulator treatment), as that variable had both low median variance explained and was correlated with baseline immunomodulator use. The sample size is large compared to the number of degrees of freedom (df) lost by including predictors that may not be relevant for some genes.

As expected, cell proportions were among the biological factors that explained the most variance on average; they are one of the largest sources of variation in bulk blood expression data, and are a major driver of transcriptional response to immune perturbations.[12] In DGE analyses, two sets of separate models were fit including and excluding cell proportions as predictors, but otherwise identical. In models without cell proportions included, differential expression after drug perturbation could represent up or downregulation on a per-cell basis, but could also come from differences in cell proportions induced by anti-TNF treatment. The estimates from models adjusted for cell proportions are more likely to reflect up or downregulation on a per-cell basis.

#### Linear contrasts for pairwise group comparisons

Per-gene DGE models were fit in dream.[13] Like the variance components analysis models, these DGE models were linear mixed models:

$$y = 0 + \beta_{G_{trd}} G_{trd} + \sum^{9} \beta_Z Z + \left( \sum^{5} \beta_C C \right) + u + v + \varepsilon$$

where:

- The response variable is normalised gene expression *y*.
- 0 indicates a group means parameterisation, where the fixed intercept term is omitted to avoid over-parameterisation.
- $G_{trd}$ is a fixed effect for experimental group defined by combinations of the predictors of interest: timepoint *t* (week 0, 14, 30, 54), response *r* (PR, PNR), and drug *d* (infliximab, adalimumab). This is equivalent to having an intercept term and a three-way interaction between visit, response, and drug, including all lower order terms, but is more convenient for testing pairwise expression differences between groups, as the coefficient for each term is the estimate of mean expression for that group.
- $\sum^{9} \beta_Z Z$ are the nine non-cell proportion fixed effects from variable selection (Fig. S2): sex (Sex), age of disease onset (Age_of_Onset), disease duration (Disease_Duration), smoking history (Smoking_History: ex, current or never), whether the patient has had surgery for CD (Crohns_Surgery), whether the patient was on immunomodulator at baseline (On_Immunomodulator_At_Baseline), whether the patient was on steroids at baseline (On_Steroids_At_Baseline), BMI at baseline (Earliest_BMI), and library preparation protocol version (Library_Prep_Protocol).
- $(\sum^{5} \beta_C C)$ are the five cell proportion fixed effects from variable selection for NK cells, monocytes, B cells, CD4+ T cells, and CD8+ T cells.
- *u* is a random intercept for RNA-seq plate (RNA_Plate).
- *v* is a random intercept for patient (PANTS.ID), nested inside RNA-seq plate.

Four sets of per-gene models were fit, with or without the cell proportion terms ($\sum^5 \beta_C C$), and replacing $\beta_{G_{trd}} G_{trd}$ (separate drug models) with $\beta_{G_{tr}} G_{tr} + \beta_d d$ (pooled drug models) or not. Unlike with variance components analysis, to avoid small-sample bias in estimates of fixed effect standard errors, REML was used for estimation.[14] Specific hypotheses were tested using sum-to-zero contrasts, which are linear combinations of model coefficients with weights summing to zero. For example, to test for DGE between responders and non-responders to infliximab at baseline in the non-pooled model, we used a contrast where the weight for the week 0/responder/infliximab group coefficient was 1, the weight for the week 0/non-responder/infliximab group coefficient was -1, and all other coefficient weights were zero. To compute p-values, the contrast divided by its standard error was compared to the $t$-distribution using the Satterthwaite approximation for df. False discovery rate (FDR) was controlled with the Benjamini-Hochberg (BH) method,[15] with threshold set at 0.05, computed separately for each contrast.

#### Spline model comparing primary responders and non-responders over time

The aim was to use expression data from all four timepoints to find genes associated with response, while avoiding a large number of pairwise comparisons. We fit a natural cubic spline (splines::ns, R v3.6.2)[16] to the study day to allow for non-linear trajectories of expression over time. We set two inner knots at week 14 (day 98) and week 30 (day 210) representing the centre of the study day windows for those two visits, as expression was expected to change after each drug dose. To include all data within the boundaries, the two boundary knots were set at the minimum and maximum values of study day rather than week 0 and week 54. A basis matrix was computed with ns(Study_Day, knots = 7*c(14, 30)), giving a matrix with three columns, each column being a transformation of study day. The columns were fit in the regression model in place of study day to allow for non-linear effects of study day on expression. The model form used was the same as that used for pairwise contrasts, except with $\beta_{G_{trd}} G_{trd}$ replaced by $\beta_r r + \sum^3 \beta_b b + \sum^3 \beta_{rb} rb + \beta_d d$, where *r* is response status, *d* is drug, $\sum^3 \beta_b b$ are the three columns of the basis matrix, and $\sum^3 \beta_{rb} rb$ are the second-order interaction terms between response status and the basis matrix columns. Separate sets of per-gene models were again fit with and without cell proportion terms ($\sum^5 \beta_C C$).

When testing for response-associated differences in the spline parameters, the predictors of interest were the interaction terms $\sum^3 \beta_{rb} rb$. The three terms were tested jointly using an F-test, with the FDR threshold set at 0.05 (BH method). A significant result indicates a significant difference in the trajectory of expression over study day between responders and non-responders.

##### Clustering expression over all timepoints

We clustered genes by their expression trajectories to define sets of genes with similar trajectories over time. This was done to aid the interpretation of significant genes from the cell proportion-adjusted spline model using gene set enrichment analysis. Expression data was converted to the CPM scale using TMM normalisation factors, then regressed against cell proportions. Residuals were centred and scaled per gene. A distance matrix was computed using 1 − *r* as the distance metric, where *r* is the Pearson correlation. Hierarchical clustering was performed with complete agglomeration for inter-cluster distance (fastcluster::hclust(method = "complete")).[17] The optimal number of clusters was assessed by the gap statistic (factoextra::fviz_nbclust(method = "gap_stat", nboot = 500)), which determines when the change in within-cluster dispersions are no longer significantly improved by increasing the number of clusters.[18] The default "firstSEmax" criteria was used to choose the optimal number of clusters *k*, which finds the first local maximum at m clusters where $Gap(m) \geq Gap(m+1)$, then finds the smallest $k\colon 1 \leq k \leq m$ such that Gap(*k*) is not less than Gap(*m*) minus the bootstrapped standard error of Gap(*m*). The hierarchical clustering tree was then cut into *k* clusters.

#### Gene set enrichment analyses

##### Ranked gene set enrichment

The gene sets used for ranked gene set enrichment analyses were blood transcription modules (BTMs) available in the tmod package.[19] Modules are "specific to the context of immune responses in blood tissue": sets of genes with transcriptional and functional similarities across a variety of healthy, diseased, and stimulated conditions.[20] The 260 modules from Chaussabel et al.[21] (prefixed "DC") were constructed by unsupervised clustering of 239 PBMC transcriptomes from multiple disease datasets, then annotated by data mining of gene names in PubMed abstracts. The 346 modules from Li et al.[20] (prefixed "LI") were constructed from coexpression analysis of approximately 30 000 blood transcriptomes, then annotated making use of Gene Ontology (GO) terms, cell type-specific markers, pathway databases, and manual literature searches. Li et al. modules are better annotated and were used as the default module set for gene set enrichment.

Ranked gene set enrichment analyses were conducted using tmod::tmodCERNOtest, which is a non-parametric test for enrichment of small ranks within specific sets of genes compared to all genes, after the genes are ranked by some metric. The CERNO statistic for a gene set is:

$$-2\sum_{i=1}^{n} ln\frac{r_i}{N} \sim \chi^2(2n)$$

where $n$ is the number of genes in the set, $N$ is the number of measured genes in the experiment, and $r_i$ is the rank of the $i$th gene in the set. For each contrast, as the t-statistics are not comparable between genes due to the use of approximate df, genes were ranked by the signed z-score reported by dream, which is a monotonic transformation of the p-value. Similarly, moderated F-statistics from the spline model are not comparable between genes, so we used the signed F-statistics computed by dream from the transformation of the p-value. FDR control for the number of gene sets tested was performed using the BH procedure, separately for each tested contrast. tmod::tmodCERNOtest is one-sided and only considers enrichment of small ranks when computing significance. As genes can be down or upregulated, separate tests were performed sorting genes in ascending and descending order, and the more significant result was used to determine the overall direction of effect for each gene set. The effect size of gene set enrichments can be quantified with the area under the curve (AUC), computed from U, the test statistic from a Mann-Whitney U test (a.k.a. Wilcoxon rank-sum test):

$$U = n(N-n) + \frac{n(n+1)}{2} - \sum_{i=1}^{n} r_i$$

Then $AUC = U/(n(N-n)) \in [0,1]$, with significant results from the one-sided tmod::tmodCERNOtest having AUC > 0.5.

##### Gene set overrepresentation

Gene set overrepresentation analyses for BTMs were run with tmod::tmodHGtest, which implements the hypergeometric test for enrichment of a query gene set within a gene module, controlling the FDR at 0.05 using the BH method. The 15 511 genes assayed by RNA-seq were used as a custom background set.

Gene set overrepresentation analyses were also run using gprofiler2::gost, which implements a similar test using publicly-available gene set databases including Gene Ontology, KEGG Reactome, WikiPathways, miRNA targets from miRTarBase, and regulatory motif matches from TRANSFAC. Multiple testing was controlled at alpha = 0.05 using the g:SCS method (correction_method = "g_SCS"), with genes assayed by RNA-seq provided as a custom background (domain_scope = "custom_annotated").[22]

#### Prediction of primary response status from expression

##### Computing module scores by single-sample GSEA

The single-sample gene set enrichment analysis method (ssGSEA 2.0, https://github.com/broadinstitute/ssGSEA2.0/, commit b3d035ed31043b277512fd4f08e90f4c1728beec) was used to compute an expression score for each module in each RNA-seq sample. This module score is a measure of coordinate up or downregulation of the genes in the module in a particular sample as compared to other samples based on expression ranks. Combining the Li et al.[20] and Chaussabel et al.[21] datasets, 427 modules had sufficient overlap with the 15 511 genes in the filtered expression matrix for ssGSEA scores to be calculated.

#### Model training and internal validation

Models were built to predict binary primary response status based on baseline expression and clinical variables. The predictors included the 427 module expression scores, as well as clinical variables (drug, sex, age of onset, disease duration, smoking status, previous CD surgery, immunomodulator use at baseline, steroid use at baseline, BMI) and cell proportions (CD4+ T cells, CD8+ T cells, B cells, NK cells, monocytes). At the week 0 timepoint, complete data was available for 152 responders and 116 non-responders; at week 14, complete data was available for 142 responders and 104 non-responders.

Model training and evaluation was performed using caret (v6.0-8).[23] Pre-processing was performed to remove zero variance predictors, then predictors were centred and scaled. To avoid lack of power during model development, no data splitting was performed (TRIPOD model type 1b).[24] The predictive methods used were: penalised and regularised logistic regression methods (plr, regLogistic, glmnet), parallel random forest (parRF), eXtreme Gradient Boosting (xgbTree), support vector machines with a radial basis (svmRadial), k-nearest neighbours (knn), naive Bayes (naive_bayes), and Gaussian process models ( gaussprLinear). To evaluate the incremental advantage of adding gene expression information, each method was trained with each of three sets of predictors. Initially only clinical variables were used as predictors, then cell proportions were added the the predictor set, and finally ssGSEA module scores were added. Model parameters were tuned by resampling (50 bootstraps per method, predictor set, and parameter combination; trainControl(method = “boot”)), optimising using the AUC metric (train(method = “ROC”). Each bootstrap involves resampling the dataset with replacement to obtain a bootstrap dataset of the same size (on average 63.2% of observations will be sampled one or more times), training a predictive model on the bootstrap dataset, then evaluating the model AUC on the observations that were not sampled (the “out-of-bag” observations, on average 36.8% of observations). The parameter combination with the highest mean AUC over 50 bootstraps was selected for the final model. The resampling metrics (AUCs) from the final model per method and predictor set were compared to identify the best performing models. Bootstrapping provides internal estimates of model performance without data splitting, which is undesirable given the relatively small sample size.[25]

A single AUC for each final tuned model was computed after merging all 50 bootstraps. ROC curves were plotted with pROC (v1.18.0).[26] Pairwise tests for differences in AUC between ROC curves were performed with pROC::roc.test(method = "bootstrap", boot.n = 2000), which uses bootstrapping to estimate the standard deviation of the difference in AUCs.

### Supplementary Results

#### Evaluating possible contributors to heterogeneity of effects between drug subgroups

We considered factors that may contribute to the heterogeneity observed for baseline models unadjusted for cell composition: 859 differentially expressed genes for infliximab patients versus only one for adalimumab patients. Power differences related to sample size were not the sole contributor. Although there was a greater sample size for the infliximab subgroup (Fig. S1a), heterogeneity remained after randomly downsampling the infliximab subgroup to the same size as the adalimumab subgroup (Fig. S5). To examine the effect of adjusting for clinical variables on the association between response and expression, a set of models was run excluding all clinical covariates, such that the only predictor terms in the model were grouping variables (timepoint, response status, drug), technical terms (library preparation protocol and plate), and the random intercept for patient. A set of models was run including additional two-way interaction terms between drug and components of the propensity score used to match IFX patients during RNA-seq sample selection: sex, age, immunomodulator usage at baseline, steroid usage at baseline, and BMI. This allows coefficient estimates for the propensity score terms to vary between drugs. These two sets of models were run per drug subgroup, adjusted or unadjusted for cell composition. Heterogeneity remained regardless of model form, with a high number of single-gene associations uniquely observed in the IFX subgroup unadjusted for cell composition (Fig. S4). We also considered adjusting for week 14 drug level due to its association with PNR,[1] but this was precluded by substantial missingness in drug level data (196/814 samples). Finally, we directly tested for differences in cell composition at baseline between study groups.

Infliximab responders had significantly lower proportions of CD4+ and CD8+ T cells, and higher proportions of granulocytes compared to infliximab non-responders at baseline (Mann-Whitney test, Bonferroni-corrected $p = 0.028$). These differences were not observed in adalimumab patients (Fig. S10). Thus associations between baseline expression of immune gene modules and post-induction primary response may be partially mediated by baseline differences in cell composition, and particularly in the infliximab subgroup. In the main manuscript, we focus on models adjusting for cell composition, which allows a more sensible interpretation for analyses where expression data for both drugs are pooled to increase sample size.

### Supplementary Tables

**Table S1. The UK Inflammatory Bowel Disease Pharmacogenetics Study Group.** All UK gastroenterologists were invited to participate in the PANTS study, which was promoted through the UK National Institute for Health Research and the British Society of Gastroenterology.

*See accompanying Supplementary Table and Figure files.*

### Supplementary Figure Legends

**Figure S1. Distribution of PANTS RNA-seq samples over study timepoints.**

a. RNA-seq samples stratified by timepoint and study group. Windows from Kennedy et al. for the four major PANTS visits are colored in grey. Samples mostly come from major visits, but a small number of LOR and exit visit samples were also included.
b. Numbers of patients with samples at multiple timepoints. Filled circles indicate the presence of at least one sample at the corresponding timepoint.

**Figure S2. Distributions of per-gene percentage of variance in expression explained by study variables.**

Variables are ordered by the median of per-gene variance explained estimates from variance components analysis. PANTS.ID = patient ID, NK = NK cell, Gran = granulocyte, Mono = monocyte, Bcell = B cell, CD4T = CD4+ T cell, CD8 = CD8+ T cell. random_numbers is a null drawn from the standard normal distribution. Variables other than random_numbers and Ever_Immunomodulator were retained for use in downstream DGE and prediction models.

**Figure S3. Baseline expression associated with primary response, unadjusted for cell composition.**

a. Volcano plots of DGE between responders (PR) and non-responders (PNR) at week 0; for infliximab (IFX), adalimumab (ADA), or with drug subgroups pooled. Annotated genes show significant associations from this study, and previously reported associations from the literature in both blood and gut biopsies. Dashed line shows significance threshold at FDR = 0.05.
b. Top gene modules differentially expressed between PR and PNR at week 0. Columns correspond to results for infliximab (IFX), adalimumab (ADA), difference between IFX and ADA (IFX − ADA i.e. drug-by-response interaction), and pooled drug analyses. The top 30

modules ranked by minimum FDR in any column are shown. Dashed lines show significance thresholds at FDR = 0.05.

**Figure S4. Baseline gene expression differences between responders and non-responders with varying model specification.**
Volcano plots of DGE between responders (PR) and non-responders (PNR) at week 0; for adalimumab (ADA) and infliximab (IFX) subgroups. Columns correspond to cell proportion adjusted and cell proportion unadjusted models with either clinical variables excluded, or additional interaction terms between drug and variables used in propensity score matching during RNA-seq sample selection. Annotated genes show significant associations from this study, and previously reported associations from the literature in both blood and gut biopsies. Dashed line shows significance threshold at FDR = 0.05.

**Figure S5. Baseline gene expression differences between responders and non-responders, with downsampling to match baseline IFX and ADA subgroup sample sizes.**
Volcano plots of DGE between responders (PR) and non-responders (PNR) at week 0; for infliximab (IFX), adalimumab (ADA), or with drug subgroups pooled. Downsampling was performed, randomly selecting 66/86 IFX PR baseline samples and 57/59 IFX PNR baseline samples to match the corresponding ADA group sizes. Annotated genes show significant associations from this study, and previously reported associations from the literature in both blood and gut biopsies. Dashed line shows significance threshold at FDR = 0.05.

**Figure S6. Top modules differentially expressed between week 14 and week 0 in a) responders (PR) and b) non-responders (PNR), adjusted for cell composition.**
Columns correspond to results for infliximab (IFX), adalimumab (ADA), difference between IFX and ADA (IFX − ADA i.e. drug-by-response interaction), and pooled drug analyses. The top 30 modules ranked by minimum FDR in any column are shown. Dashed lines show significance thresholds at FDR = 0.05.

**Figure S7. Distributions of model specificity, sensitivity and AUC (ROC) from resampling (50 bootstraps) for each combination of algorithm and predictor set at baseline.**
Metrics shown are the resampling metrics from the final tuned models. The prefixes “clinical”, “cellProps”, and “moduleScores” correspond to the inclusion of clinical variables, cell proportion estimates, and expression module scores in the predictor datasets respectively.

Suffixes indicate the method used (R 3.6, caret package).

**Figure S8. Distributions of model specificity, sensitivity and AUC (ROC) from resampling (50 bootstraps) for each combination of algorithm and predictor set at week 14.**

Metrics shown are the resampling metrics from the final tuned models. The prefixes “clinical”, “cellProps”, and “moduleScores” correspond to the inclusion of clinical variables, cell proportion estimates, and expression module scores in the predictor datasets respectively.

Suffixes indicate the method used (R 3.6, caret package).

**Figure S9. Correlation matrix of variables measured in PANTS that were considered as potential predictor variables for DGE analyses.**

NK = NK cell, Gran = granulocyte, Mono = monocyte, Bcell = B cell, CD4T = CD4+ T cell, CD8 = CD8+ T cell.

**Figure S10. Differences in baseline cell proportion estimates between study groups.**

NK = NK cell, Gran = granulocyte, Mono = monocyte, Bcell = B cell, CD4T = CD4+ T cell, CD8 = CD8+ T cell. Nominal significance of pairwise comparisons using the Mann-Whitney test (R 3.6, wilcox.test(paired=F)) is shown: ns 0.05-1, * 0.01-0.05, ** 0.001-0.01, *** 0.0001-0.001, **** <0.0001.

### Supplementary Figures

*See accompanying Supplementary Table and Figure files.*
