## Supplementary Tables and Figures for "Baseline expression of immune gene modules in blood is associated with primary response to anti-TNF therapy in Crohn’s disease patients": table_s1.docx

| **Hospital or Trust name** | **City** | **Name** | **Job Title** |
| --- | --- | --- | --- |
| Tameside Hospital NHS Foundation Trust | Ashton U Lyne | Dr Vinod Patel | Consultant Gastroenterologist |
| Basildon and Thurrock University Hospitals NHS Foundation Trust | Basildon | Dr Zia Mazhar | Consultant Gastroenterologist |
| Hampshire Hospitals NHS Foundation Trust | Basingstoke | Dr Rebecca Saich | Consultant Gastroenterologist |
| Royal United Hospital | Bath | Dr Ben Colleypriest | Consultant Gastroenterologist |
| Ulster Hospital | Belfast | Dr Tony C Tham | Consultant Gastroenterologist |
| University Hospital's Birmingham NHS Foundation Trust | Birmingham | Dr Tariq H Iqbal | Consultant Gastroenterologist |
| East Lancashire NHS Teaching Trust | Blackburn | Dr Vishal Kaushik | Consultant Gastroenterologist |
| Blackpool Teaching Hospitals NHS Foundation Trust | Blackpool | Dr Senthil Murugesan | Consultant Gastroenterologist |
| Bolton NHS Trust | Bolton | Dr Salil Singh | Consultant Gastroenterologist |
| Royal Bournemouth Hospital | Bournemouth | Dr Sean Weaver | Consultant Gastroenterologist |
| Bradford Teaching Hospitals Foundation Trust - (St Lukes Hospital &Bradford Royal Infirmary) | Bradford | Dr Cathryn Preston | Consultant Gastroenterologist |
| Brighton and Sussex University Hospitals NHS Trust | Brighton | Dr Assad Butt | Paediatric Consultant Gastroenterologist |
| Brighton and Sussex University Hospitals NHS Trust | Brighton | Dr Melissa Smith | Consultant Gastroenterologist |
| University Hospitals Bristol NHS Foundation Trust | Bristol | Dr Dharamveer Basude | Consultant Paediatric Gastroenterologist |
| University Hospitals Bristol NHS Foundation Trust | Bristol | Dr Amanda Beale | Consultant Gastroenterologist |
| Frimley Park Hospital NHS Foundation Trust | Camberley | Dr Sarah Langlands | Consultant Gastroenterologist |
| Frimley Park Hospital NHS Foundation Trust | Camberley | Dr Natalie Direkze | Consultant gastroenterologist |
| Cambridge University Hospitals NHS Foundation Trust | Cambridge | Dr Miles Parkes | Consultant Gastroenterologist |
| Cambridge University Hospitals NHS Foundation Trust | Cambridge | Dr Franco Torrente | Consultant Paediatric Gastroenterologist |
| Cambridge University Hospitals NHS Foundation Trust | Cambridge | Dr Juan De La Revella Negro | Research fellow |
| North Cumbria University Hospitals NHS Trust | Carlisle | Dr Chris Ewen MacDonald | Consultant Gastroenterologist |
| Ashford & St Peter's Hospitals NHS Foundation Trust | Chertsey | Dr Stephen M Evans | Consultant Gastroenterologist |
| St Peter's Hospital | Chertsey | Dr Anton V J Gunasekera | Consultant Gastroenterologist |
| Ashford & St Peter's Hospitals NHS Foundation Trust | Chertsey | Dr Alka Thakur | Paediatric Consultant |
| Chesterfield Royal NHS Foundation Trust | Chesterfield | Dr David Elphick | Consultant Gastroenterologist |
| Colchester Hospital University NHS Foundation Trust | Colchester | Dr Achuth Shenoy | Consultant Gastroenterologist |
| University Hospitals Coventry and Warwickshire NHS Trust | Coventry | Prof Chuka U Nwokolo | Consultant Gastroenterologist |
| County Durham and Darlington NHS Foundation Trust | Darlington | Dr Anjan Dhar | Consultant Gastroenterologist & Hon. Clinical Lecturer |
| Derby Hospital NHS Foundation NHS Trust | Derby | Dr Andrew T Cole | Consultant Gastroenterologist |
| Doncaster and Bassetlaw Hospitals NHS Foundation Trust | Doncaster | Dr Anurag Agrawal | Consultant Gastroenterologist |
| Dorset County Hospital NHS Foundation Trust | Dorchester | Dr Stephen Bridger | Consultant Gastroenterologist |
| Dorset County Hospitals Foundation Trust | Dorchester | Dr Julie Doherty | Paediatric Consultant |
| Dudley Group NHS Foundation Trust | Dudley | Dr Sheldon C Cooper | Consultant Gastroenterologist |
| Russells Hall Hospital, The Dudley Group NHS Foundation Trust | Dudley | Dr Shanika de Silva | Consultant Gastroenterologist |
| Ninewells Hospital & Medical School | Dundee | Dr Craig Mowat | Consultant Gastroenterologist |
| East Sussex Healthcare Trust | Eastborne | Dr Phillip Mayhead | Consultant Gastroenterologist |
| NHS Lothian | Edinburgh | Dr Charlie Lees | Consultant Gastroenterologist and Honorary Senior Lecturer |
| NHS Lothian | Edinburgh | Dr Gareth Jones | Research fellow |
| Royal Devon and Exeter NHS Foundation Trust | Exeter | Dr Tariq Ahmad | Consultant Gastroenterologist |
| Royal Devon and Exeter NHS Foundation Trust | Exeter | Dr James W Hart | Consultant Paediatrician |
| Glasgow Royal Infirmary | Glasgow | Dr Daniel R Gaya | Consultant Gastroenterologist |
| Royal Hospital for Children | Glasgow | Prof Richard K Russell | Consultant Paediatric Gastroenterologist |
| Royal Hospital for Children | Glasgow | Dr Lisa Gervais | Research fellow |
| Gloucestershire Hospitals NHS Trust | Gloucester | Dr Paul Dunckley | Consultant Gastroneterologist |
| United Lincolnshire Hospitals NHS Trust | Grantham | Dr Tariq Mahmood | Consultant Gastroenterologist |
| James Paget University Hospitals NHS Foundation Trust | Great Yarmouth | Dr Paul J R Banim | Consultant Gastroneterologist |
| Calderdale and Huddersfield NHS Trust | Halifax | Dr Sunil Sonwalkar | Consultant Gastroenterologist |
| Princess Alexandra Hospital NHS Trust | Harlow | Dr Deb Ghosh | Consultant Gastroenterologist |
| Princess Alexandra Hospital NHS Trust | Harlow | Dr Rosemary H Phillips | Consultant Gastroenterologist |
| Hull and East Yorkshire NHS Trust | Hull | Dr Amer Azaz | Paediatric Consultant Gastroenterologist |
| Hull and East Yorkshire NHS Trust | Hull | Dr Shaji Sebastian | Consultant Gastroenterologist |
| Airedale NHS Foundation Trust | Keighley | Dr Richard Shenderey | Consultant Gastroenterologist |
| Crosshouse Hospital | Kilmarnock | Dr Lawrence Armstrong | Consultant Paediatrician |
| Crosshouse Hospital | Kilmarnock | Dr Claire Bell | Research fellow |
| The Queen Elizabeth Hospital NHS Foundation Trust | Kings Lynn | Dr Radhakrishnan Hariraj | Consultant Gastroenterologist |
| Kingston Hospital NHS Trust | Kingston upon Thames | Dr Helen Matthews | Consultant Gastroenterologist |
| NHS Fife | Kirkcaldy | Dr Hasnain Jafferbhoy | Consultant Gastroenterologist |
| Leeds Teaching Hospitals NHS Trust | Leeds | Dr Christian P Selinger | Consultant Gastroenterologist |
| Leeds Teaching Hospitals NHS Trust | Leeds | Dr Veena Zamvar | Paediatric Consultant Gastroenteorlogist |
| University Hospitals of Leicester NHS Trust | Leicester | Prof John S De Caestecker | Consultant Gastroenterologist |
| University Hospitals of Leicester NHS Trust | Leicester | Dr Anne Willmott | Paediatric Consultant Gastroenterologist |
| Mid Cheshire Hospitals NHS Foundation Trust | Leighton | Mr Richard Miller | Research Nurse |
| United Lincolnshire Hospitals NHS Trust | Lincoln | Dr Palani Sathish Babu | Consultant Gastroenterologist |
| Alder Hey Childrens Hospital | Liverpool | Dr Christos Tzivinikos | Consultant Paediatric Gastroenterologist |
| University College London Hospitals NHS Foundation Trust | London | Dr Stuart L Bloom | Consultant Gastroenterologist |
| Kings College Hospital NHS Foundation Trust | London | Dr Guy Chung-Faye | Consultant Gastroenterologist |
| Royal London Childrens Hospital, Barts Health NHS Trust | London | Prof Nicholas M Croft | Paediatric Consultant Gastroenterologist |
| Chelsea & Westminster Hospital | London | Dr John ME Fell | Consultant Paediatric Gastroenterologist |
| Chelsea and Westminster Hospital NHS Foundation | London | Dr Marcus Harbord | Consultant Gastroenterologist |
| North West London Hospitals NHS Trust | London | Dr Ailsa Hart | Consultant Gastroenterologist |
| Kings College Hospital NHS Foundation Trust | London | Dr Ben Hope | Consultant Paediatrician |
| Guys & St Thomas' NHS Foundation Trust | London | Dr Peter M Irving | Consultant Gastroenterologist |
| Barts and The London NHS Trust | London | Prof James O Lindsay | Consultant Gastroenterologist |
| Guy's and St Thomas' NHS trust | London | Dr Joel E Mawdsley | Gastroenterology Consultant |
| Lewisham and Greenwich Healthcare NHS Trust | London | Dr Alistair McNair | Consultant Gastroenterologist |
| Chelsea and Westminster Hospital NHS Foundation | London | Dr Kevin J Monahan | Consultant Gastroenterologist |
| Royal Free London NHS Foundation Trust | London | Dr Charles D Murray | Consultant Gastroenterologist |
| Imperial College Healthcare NHS Trust | London | Prof Timothy Orchard | Consultant Gastroenterologist |
| St George's Healthcare NHS Trust | London | Dr Thankam Paul | Paediatric Consultant Gastroenterologist |
| St George's Healthcare NHS Trust | London | Dr Richard Pollok | Reader and Consultant Gastroenterologist |
| Great Ormond Street Hospital for Children NHS Foundation Trust | London | Dr Neil Shah | Consultant Gastroenterologist |
| North West London Hospitals NHS Trust | London | Dr Sonia Bouri | Research fellow |
| The Luton & Dunstable University Hospital | Luton | Dr Matt W Johnson | Consultant Gastroenterologist |
| Luton and Dunstable Hospital Foundation Trust | Luton | Dr Anita Modi | Paediatric Consultant with Allergy and Gastroenterology interest |
| The Luton & Dunstable University Hospital | Luton | Dr Kasamu Dawa Kabiru | Research fellow |
| Maidstone and Tunbridge Wells NHS Trust | Maidstone | Dr B K Baburajan | Consultant Gastroenterologist |
| Maidstone and Tunbridge Wells NHS Trust | Maidstone | Prof Bim Bhaduri | Paediatric Consultant Gastroenterologist |
| Manchester University Hospitals NHS Foundation Trust | Manchester | Dr Andrew Adebayo Fagbemi | Consultant Gastroenterologist |
| Central Manchester University Hospitals NHS Foundation Trust | Manchester | Dr Scott Levison | Consultant Gastroenterologist |
| The Pennine Acute Hospitals NHS Trust | Manchester | Dr Jimmy K Limdi | Consultant Gastroenterologist |
| Manchester University NHS Foundation Trust, Wythenshawe Hospital | Manchester | Dr Gill Watts | Consultant Gastroenterologist |
| Sherwood Forest Hospitals NHS Foundation Trust | Mansfield | Dr Stephen Foley | Consultant Gastroenterologist |
| South Tees Hospital NHS Foundation Trust | Middlesbrough | Dr Arvind Ramadas | Consultant Gastroenterologist |
| Milton Keynes Hospital NHS Foundation Trust | Milton Keynes | Dr George MacFaul | Consultant Gastroenterologist |
| Newcastle Upon Tyne Hospital Trust | Newcastle | Dr John Mansfield | Consultant Gastroenterologist |
| Isle of Wight NHS Foundation Trust | Newport | Dr Leonie Grellier | Consultant Gastroenterologist |
| Norfolk & Norwich University Hospital NHS Foundation Trust | Norwich | Dr Mary-Anne Morris | Consultant Paediatric Gastroenterologist |
| Norfolk & Norwich University Hospital NHS Foundation Trust | Norwich | Dr Mark Tremelling | Consultant Gastroenterologist |
| Nottingham University Hospitals NHS Trust | Nottingham | Prof Chris Hawkey | Consultant Gastroenterologist |
| Nottingham University Hospitals NHS Trust | Nottingham | Dr Sian Kirkham | Consultant Paediatric Gastroenterologist |
| Nottingham University Hospitals NHS Trust | Nottingham | Dr Charles PJ Charlton | Consultant gastroenterologist |
| Oxford University Hospitals NHS Foundation Trust | Oxford | Dr Astor Rodrigues | Paediatric Consultant Gastroenterologist |
| Oxford University Hospitals NHS Trust | Oxford | Prof Alison Simmons | Consultant Gastroenterologist |
| Plymouth Hospitals NHS Trust | Plymouth | Dr Stephen J Lewis | Consultant Gastroenterologist |
| Poole Hospital NHS Foundation Trust | Poole | Dr Jonathon Snook | Consultant Gastroenterologist |
| Poole Hospital NHS Foundation Trust | Poole | Dr Mark Tighe | Paediatric Consultant with interest in Oncology and Gastroenterology |
| Portsmouth Hospitals NHS Trust | Portsmouth | Dr Patrick M Goggin | Consultant Gastroenterologist |
| Royal Berkshire NHS Foundation Trust | Reading | Dr Aminda N De Silva | Consultant Gastroenterologist |
| Salford Royal NHS Foundation Trust | Salford | Prof Simon Lal | Consultant Gastroenterologist |
| Shrewsbury and Telford Hospital NHS Trust | Shrewsbury | Dr Mark S Smith | Consultant Gastroenterologist |
| South Tyneside NHS Foundation Trust | South Shields | Dr Simon Panter | Consultant Gastroenterologist |
| Southampton University Hospitals NHS Trust | Southampton | Dr Fraser Cummings | Consultant Gastroenterologist |
| Southampton University Hospitals NHS Trust | Southampton | Dr Suranga Dharmisari | Research fellow |
| East and North Herts NHS Trust | Stevenage | Dr Martyn Carter | Consultant Gastroenterologist |
| NHS Forth Valley | Stirling | Dr David Watts | Consultant Gastroenterologist |
| Stockport NHS foundation Trust | Stockport | Dr Zahid Mahmood | Consultant Gastroenterologist |
| North Tees and Hartlepool NHS Foundation Trust | Stockton | Dr Bruce McLain | Paediatric Consultant Gastroenterologist |
| University Hospitals of North Staffordshire | Stoke-on Trent | Dr Sandip Sen | Consultant Gastroenterologist |
| University Hospitals of North Midlands NHS Trust | Stoke-on-Trent | Dr Anna J Pigott | Consultant Paediatric Gastroenterologist |
| City Hospitals Sunderland NHS Foundation Trust | Sunderland | Dr David Hobday | Consultant Gastroenterologist |
| Taunton and Somerset NHS Foundation Trust | Taunton | Dr Emma Wesley | Consultant Gastroenterologist |
| South Devon Healthcare NHS Foundation Trust | Torquay | Dr Richard Johnston | Consultant Gastroenterologist |
| South Devon Healthcare NHS Foundation Trust | Torquay | Dr Cathryn Edwards | Consultant gastroenterologist |
| Royal Cornwall Hospitals NHS Trust | Truro | Dr John Beckly | Consultant Gastroenterologist |
| Mid Yorkshire Hospitals NHS Trust | Wakefield | Dr Deven Vani | Consultant Physician & Gastroenterologist |
| Warrington& Halton NHS Foundation | Warrington | Dr Subramaniam Ramakrishnan | Consultant Gastroenterologist |
| West Hertfordshire Hospitals NHS Trust | Watford | Dr Rakesh Chaudhary | Consultant Gastroenterologist |
| Sandwell and West Birmingham Hospitals NHS Trust | West Bromwich | Dr Nigel J Trudgill | Consultant Gastroenterologist |
| Sandwell and West Birmingham Hospitals NHS Trust | West Bromwich | Dr Rachel Cooney | Consultant gastroenterologist |
| Weston Area Health NHS Trust | Weston-Super-Mare | Dr Andy Bell | Consultant Gastroenterologist |
| Royal Albert Edward Infirmary, Wrightington, Wigan & Leigh NHS Foundation Trust | Wigan | Dr Neeraj Prasad | Consultant Gastroenterologist |
| Hampshire Hospitals NHS Foundation Trust | Winchester | Dr John N Gordon | Consultant Gastroenterologist |
| Royal Wolverhampton Hospitals NHS Trust | Wolverhampton | Prof Matthew J Brookes | Consultant Gastroenterologist |
| Western Sussex Hospitals NHS Trust | Worthing | Dr Andy Li | Consultant Gastroenterologist |
| Yeovil District Hospital NHS Foundation Trust | Yeovil | Dr Stephen Gore | Consultant Gastroenterologist |
