## Supplementary figures and images for "Baseline expression of immune gene modules in blood is associated with primary response to anti-TNF therapy in Crohn’s disease patients"

### fig_s1.png

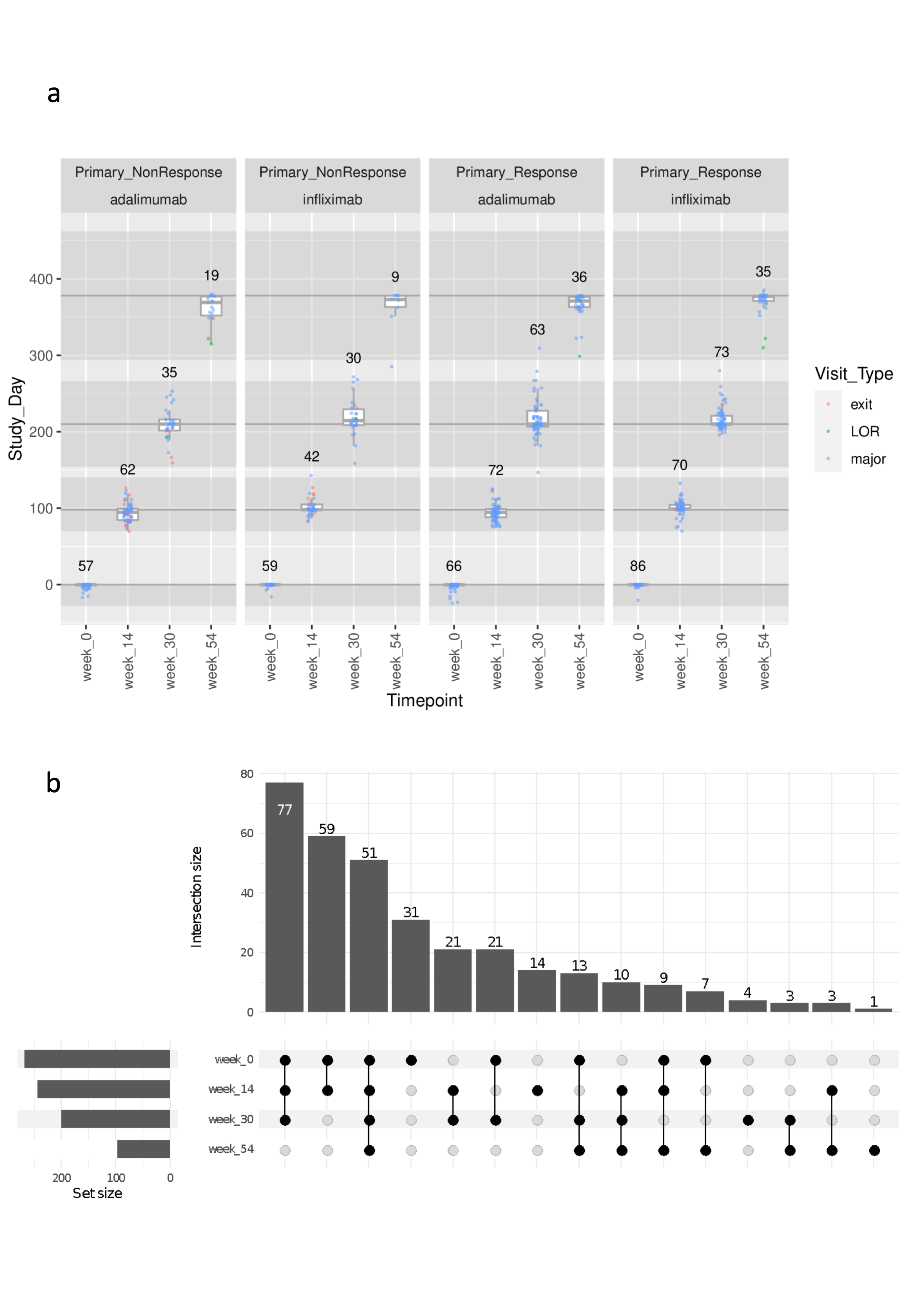

### fig_s2.png

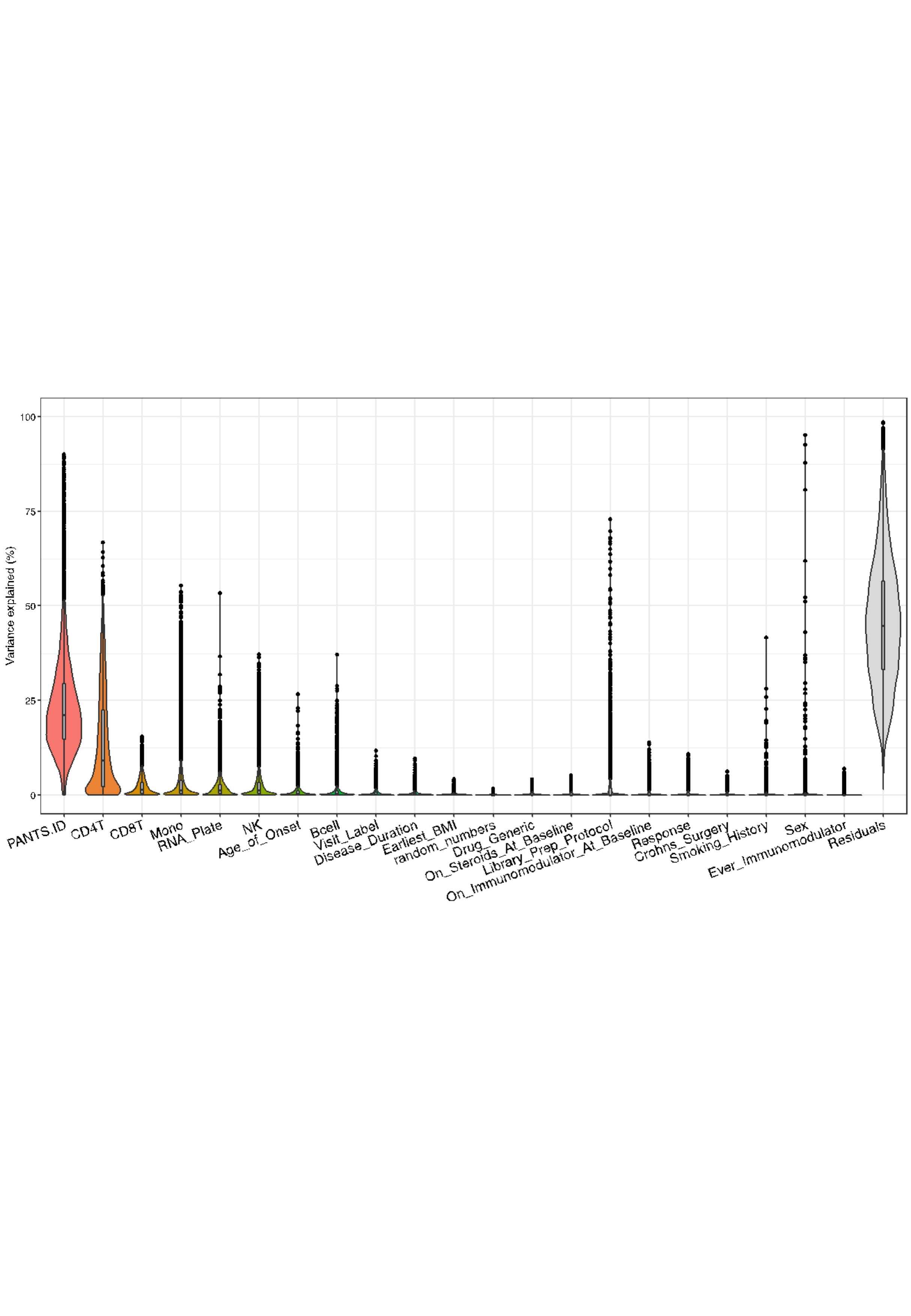

### fig_s3.png

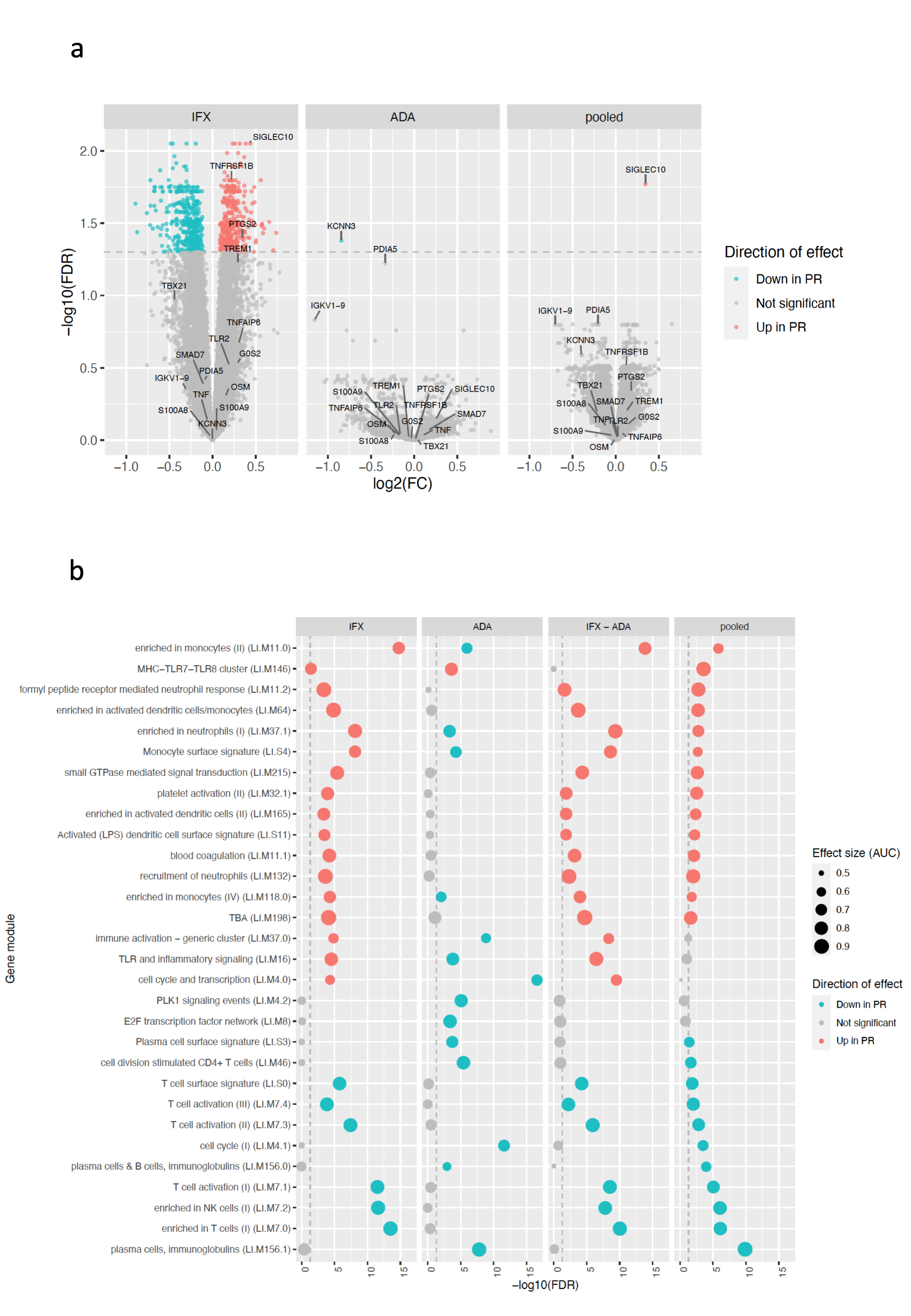

### fig_s4.png

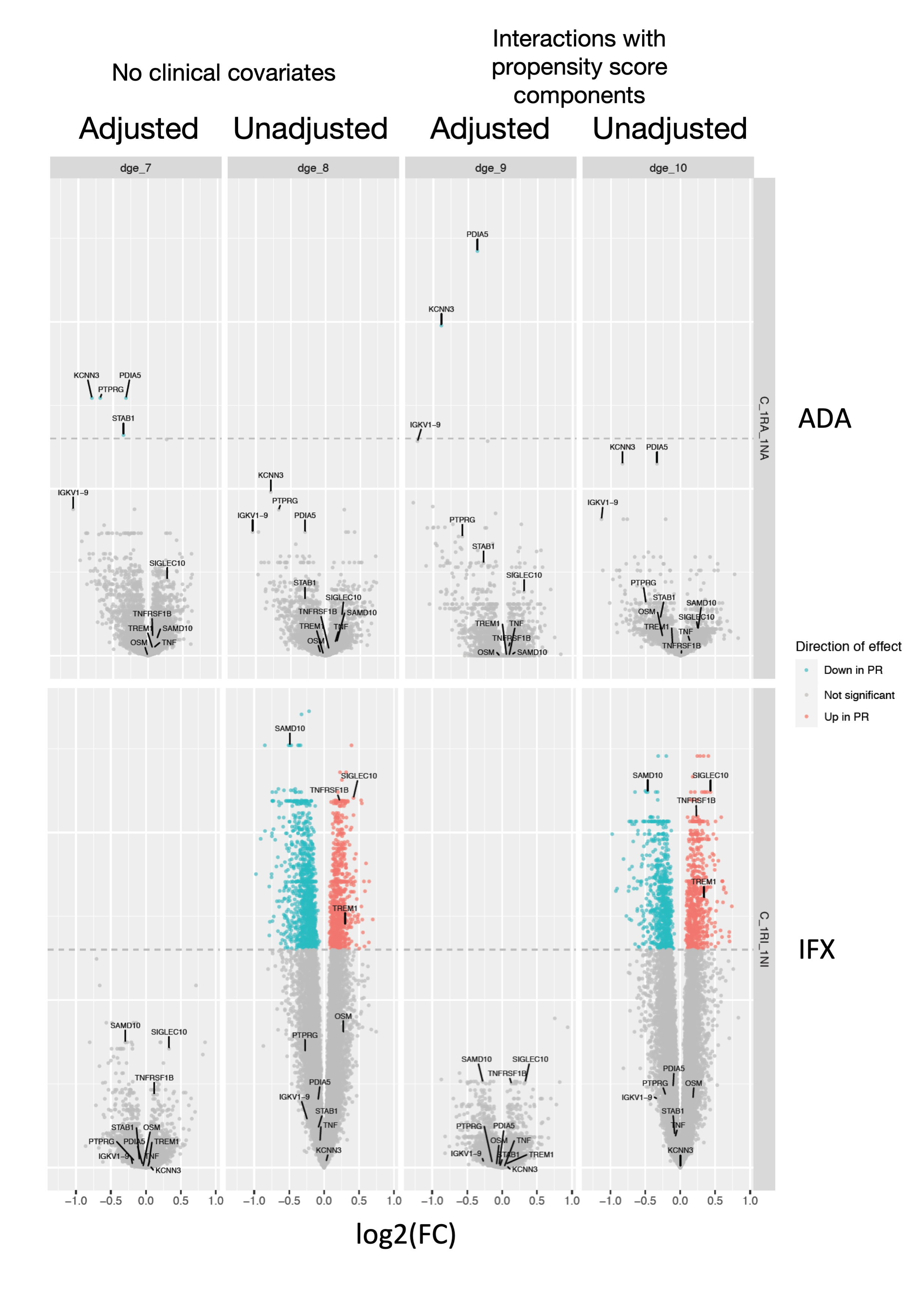

### fig_s5.png

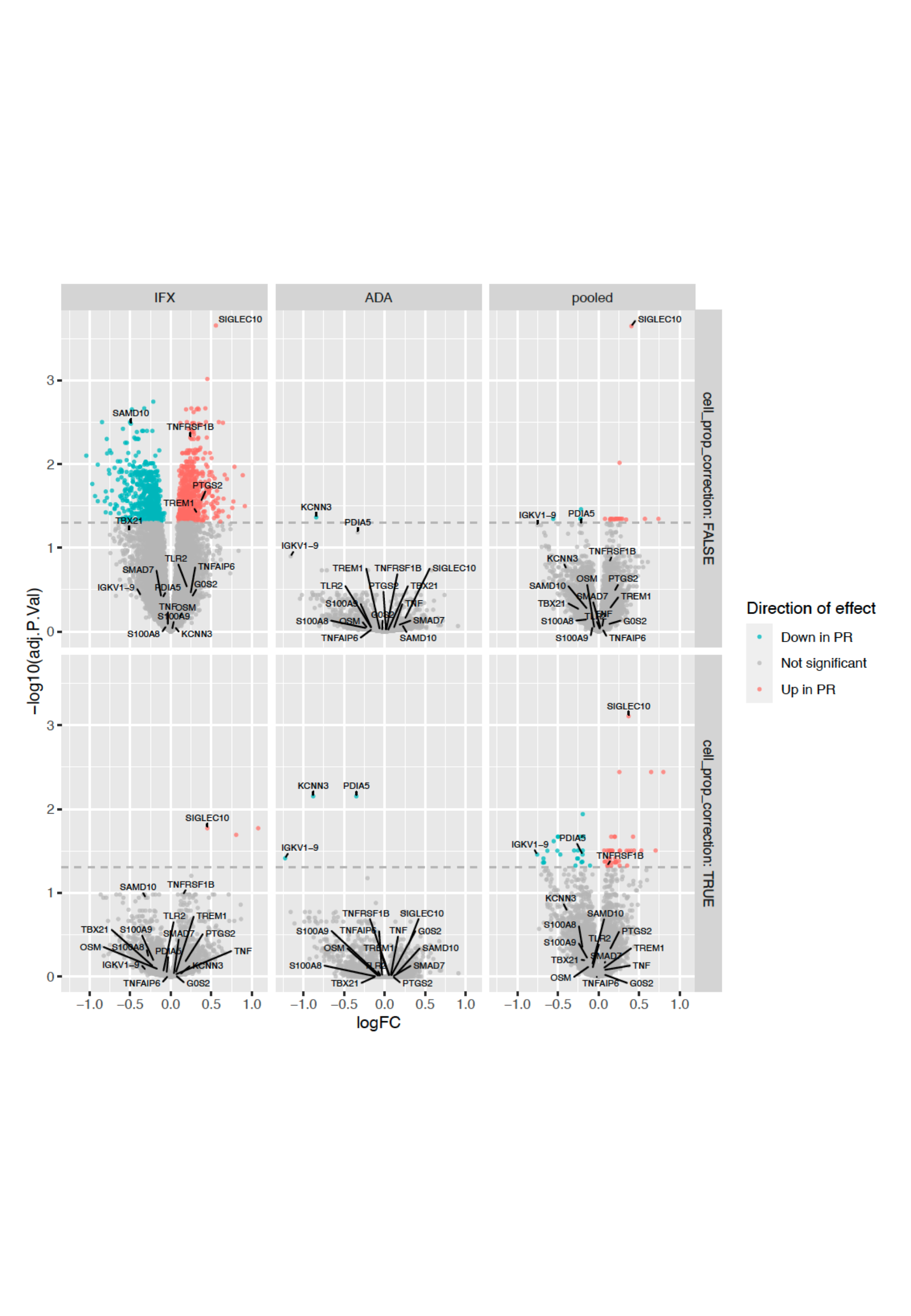

### fig_s6.png

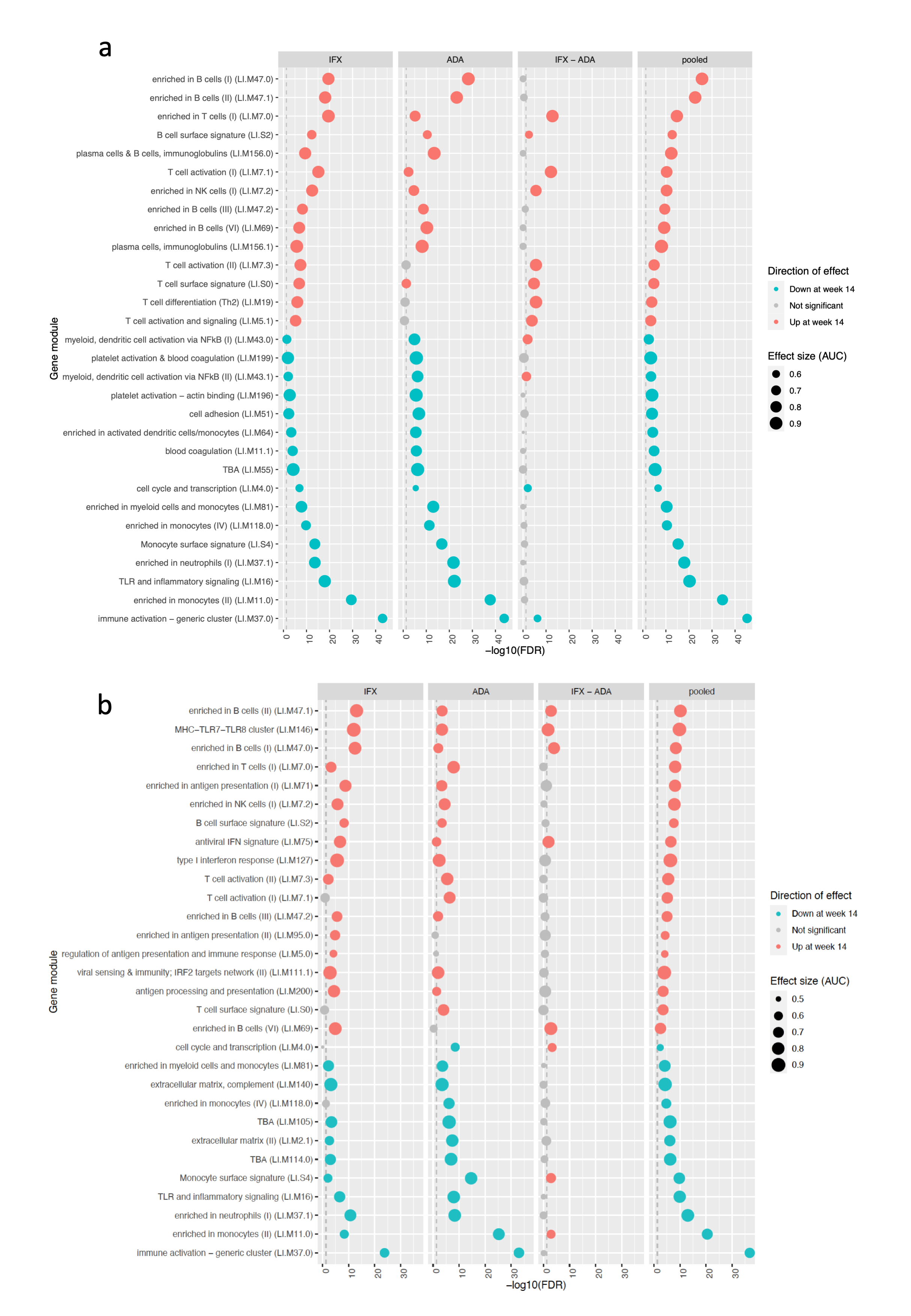

### fig_s7.png

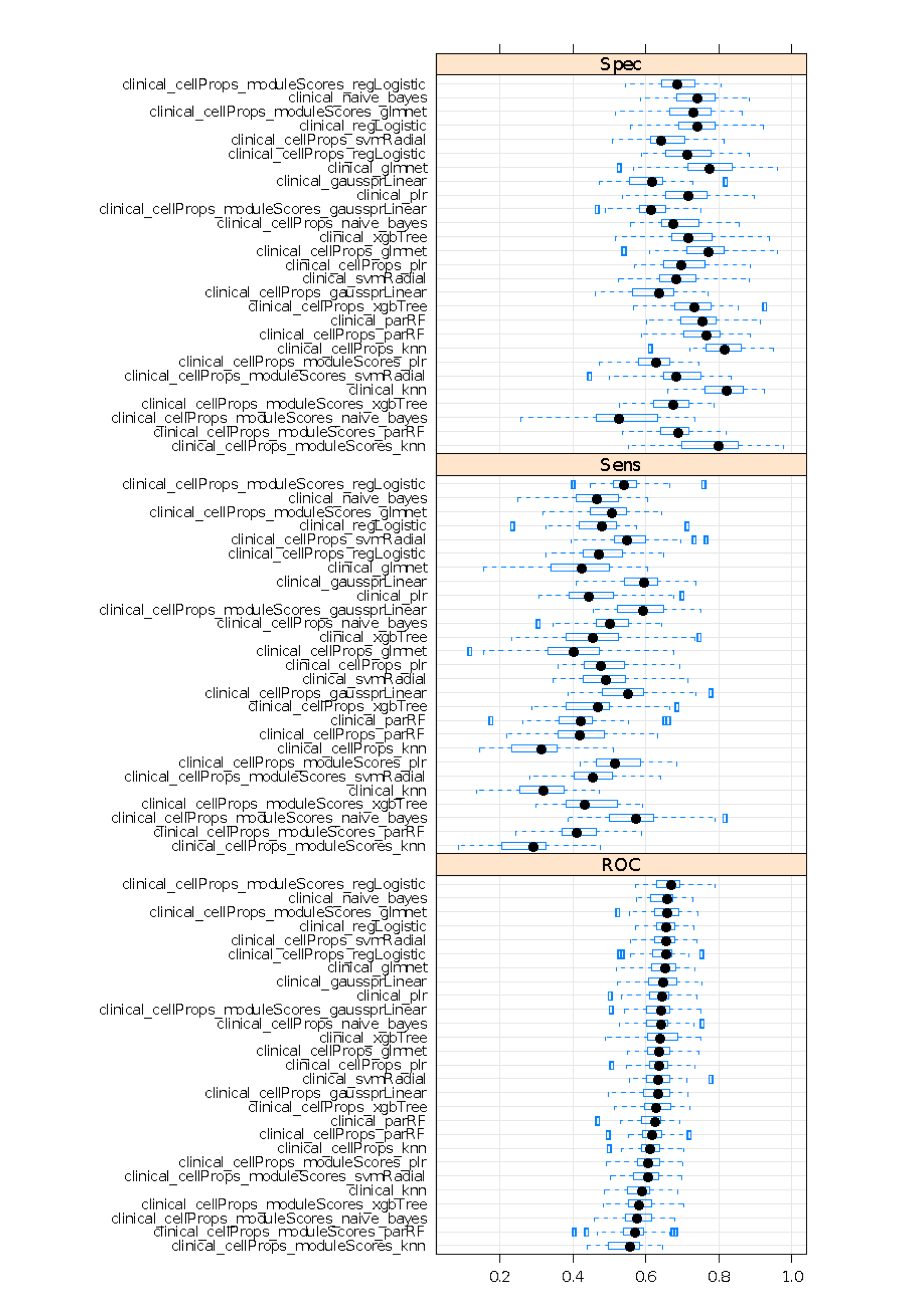

### fig_s8.png

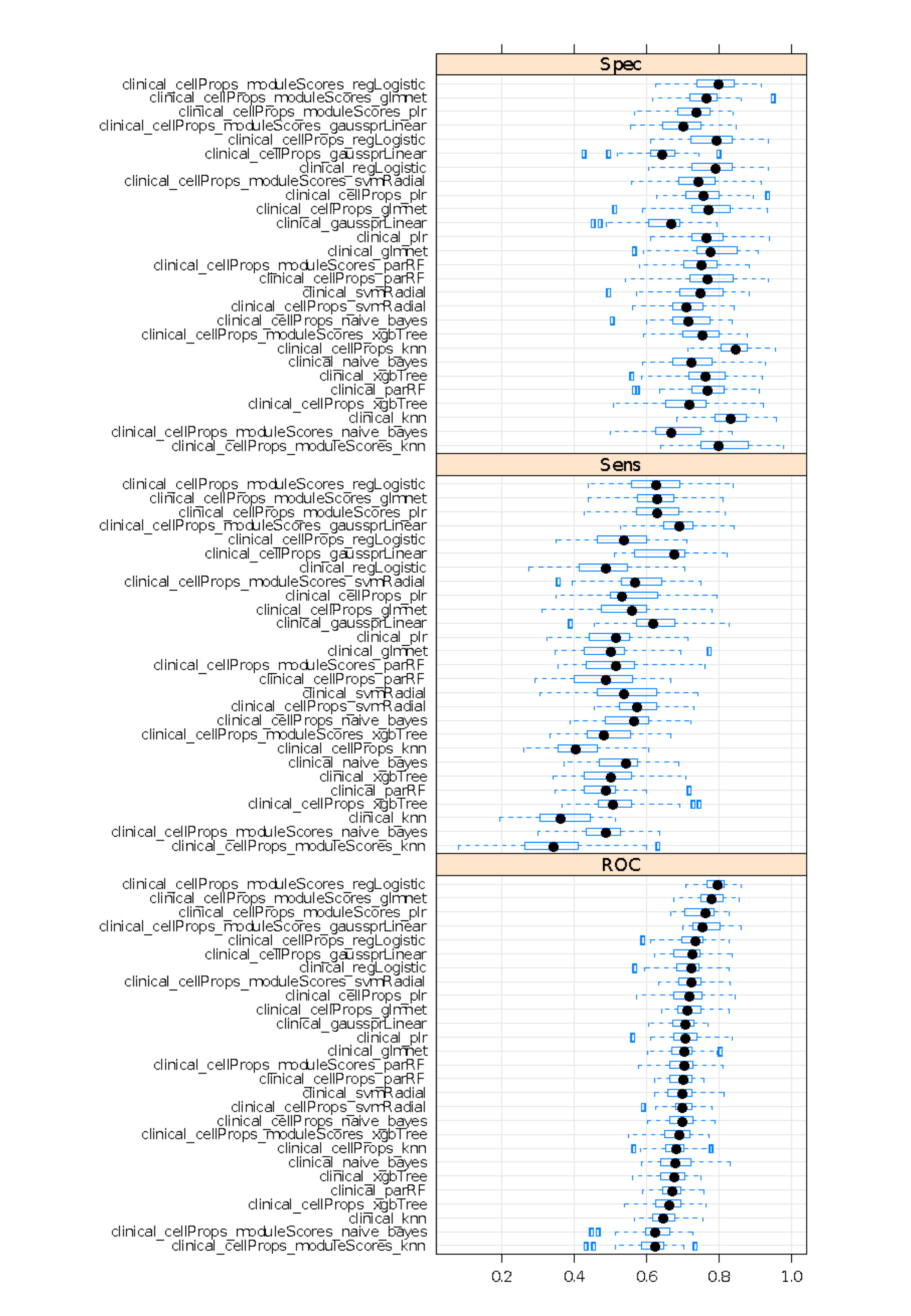

### fig_s9.png

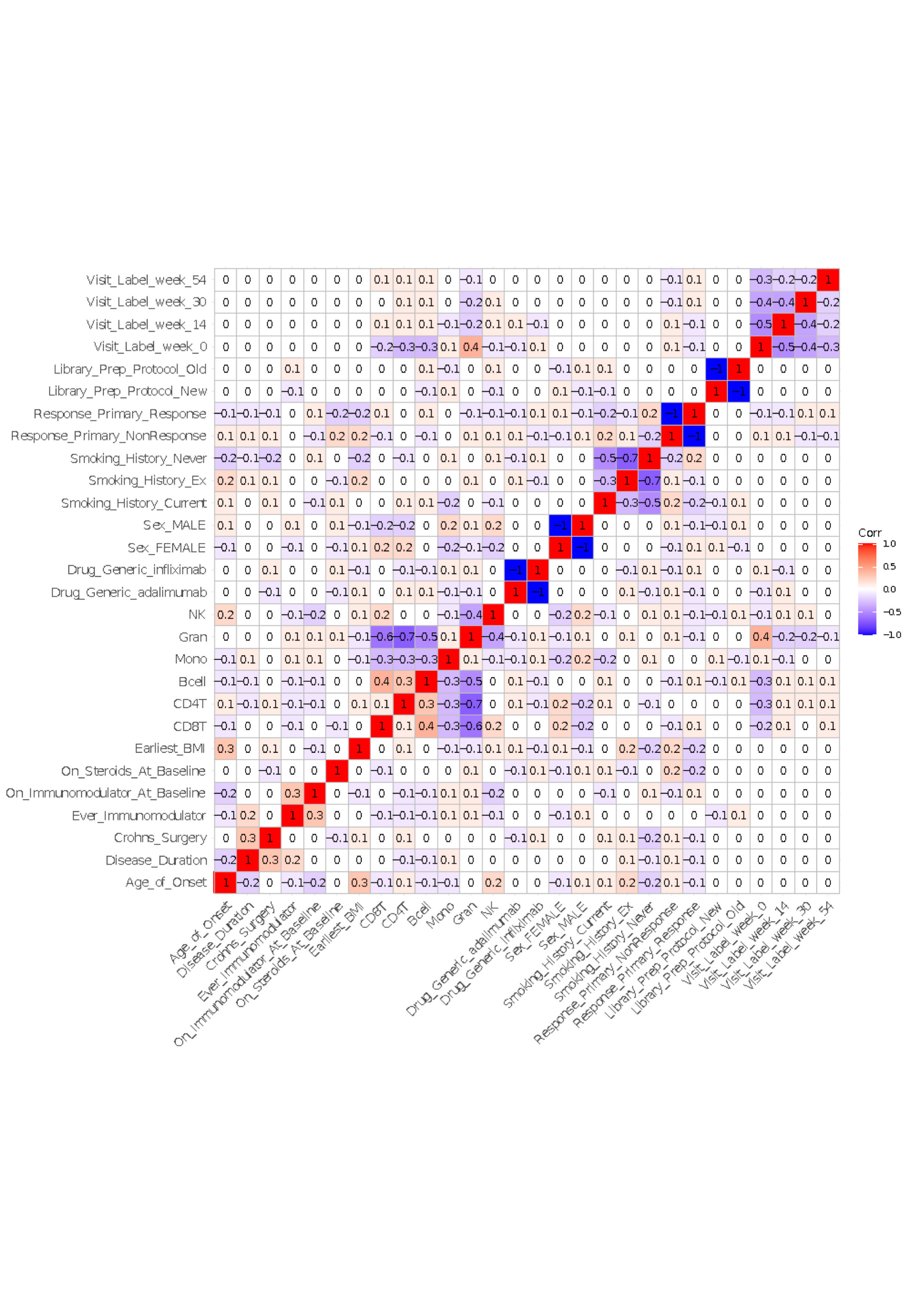

### fig_s10.png

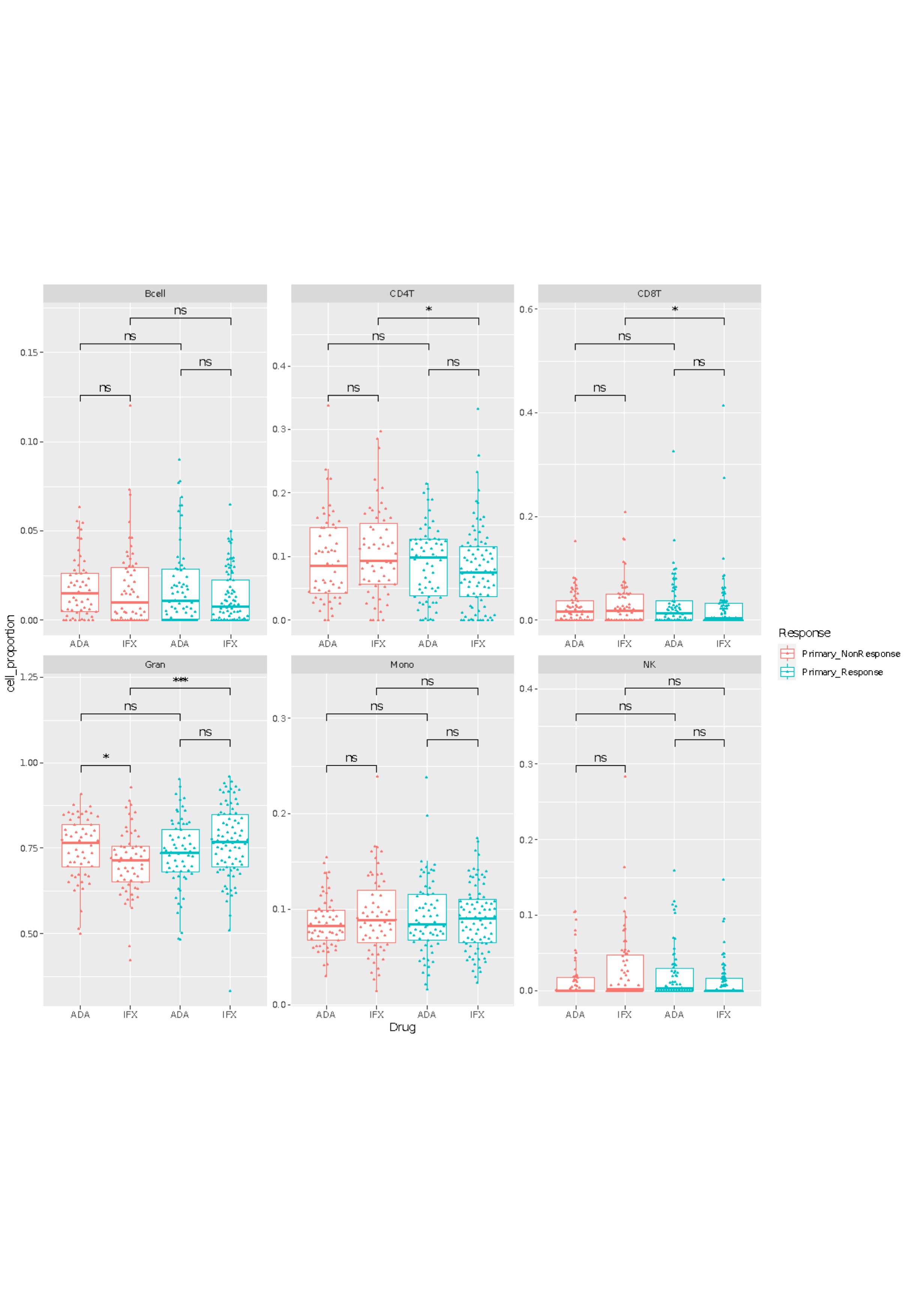
