## Supplementary material for "Baseline expression of immune gene modules in blood is associated with primary response to anti-TNF therapy in Crohn’s disease patients": README.txt

Descriptions of supplementary data files.


Data associated with
├── fig_1a

    Per-gene summary statistics from DGE model: week 0 PR IFX vs. week 0 PNR IFX, adjusted for cell composition.
│   ├── dream.gene_expr_filtered_vobj_fit_dge_1.topTable_C_1RI_1NI.csv
    Per-gene summary statistics from DGE model: week 0 PR ADA vs. week 0 PNR ADA, adjusted for cell composition.
│   ├── dream.gene_expr_filtered_vobj_fit_dge_1.topTable_C_1RA_1NA.csv
    Per-gene summary statistics from DGE model: week 0 PR vs. week 0 PNR, adjusted for cell composition.
│   └── dream.gene_expr_filtered_vobj_fit_dge_5.topTable_C_1R_1N.csv


Data associated with
├── fig_1b

Ranked gene set enrichment results, ranking genes by signed z-score from per-gene DGE models (week 0 PR IFX vs. week 0 PNR IFX, adjusted for cell composition) in ascending order to test for downregulated gene sets.
│   ├── dream.gene_expr_filtered_vobj_fit_dge_1.topTable_C_1RI_1NI.gene_set_enrichment.tmodCERNOtest_result_LI_ascending.csv
Ranked gene set enrichment results, ranking genes by signed z-score from per-gene DGE models (week 0 PR IFX vs. week 0 PNR IFX, adjusted for cell composition) in descending order to test for upregulated gene sets.
│   ├── dream.gene_expr_filtered_vobj_fit_dge_1.topTable_C_1RI_1NI.gene_set_enrichment.tmodCERNOtest_result_LI_descending.csv

Ranked gene set enrichment results, ranking genes by signed z-score from per-gene DGE models (week 0 PR ADA vs. week 0 PNR ADA, adjusted for cell composition) in ascending order to test for downregulated gene sets.
│   ├── dream.gene_expr_filtered_vobj_fit_dge_1.topTable_C_1RA_1NA.gene_set_enrichment.tmodCERNOtest_result_LI_ascending.csv
Ranked gene set enrichment results, ranking genes by signed z-score from per-gene DGE models (week 0 PR ADA vs. week 0 PNR ADA, adjusted for cell composition) in descending order to test for upregulated gene sets.
│   ├── dream.gene_expr_filtered_vobj_fit_dge_1.topTable_C_1RA_1NA.gene_set_enrichment.tmodCERNOtest_result_LI_descending.csv

Ranked gene set enrichment results, ranking genes by signed z-score from per-gene DGE models (week 0 PR/PNR x IFX/ADA interaction, adjusted for cell composition) in ascending order to test for downregulated gene sets.
│   ├── dream.gene_expr_filtered_vobj_fit_dge_1.topTable_C_(1RI_1NI)_(1RA_1NA).gene_set_enrichment.tmodCERNOtest_result_LI_ascending.csv
Ranked gene set enrichment results, ranking genes by signed z-score from per-gene DGE models (week 0 PR/PNR x IFX/ADA interaction, adjusted for cell composition) in descending order to test for upregulated gene sets.
│   ├── dream.gene_expr_filtered_vobj_fit_dge_1.topTable_C_(1RI_1NI)_(1RA_1NA).gene_set_enrichment.tmodCERNOtest_result_LI_descending.csv

Ranked gene set enrichment results, ranking genes by signed z-score from per-gene DGE models (week 0 PR vs. week 0 PNR, adjusted for cell composition) in ascending order to test for downregulated gene sets.
│   ├── dream.gene_expr_filtered_vobj_fit_dge_5.topTable_C_1R_1N.gene_set_enrichment.tmodCERNOtest_result_LI_ascending.csv
Ranked gene set enrichment results, ranking genes by signed z-score from per-gene DGE models (week 0 PR vs. week 0 PNR, adjusted for cell composition) in descending order to test for upregulated gene sets.
│   └── dream.gene_expr_filtered_vobj_fit_dge_5.topTable_C_1R_1N.gene_set_enrichment.tmodCERNOtest_result_LI_descending.csv


Data associated with
├── fig_2a

Per-gene summary statistics from DGE model: genes significant for week 14 PR vs. week 0 PR, week 14 PNR vs. week 0 PNR, and week 14/week 0 x PR/PNR interaction.
│   └── plot_gene_set_enrichment.logFC_C_3R_1R_vs_logFC_C_3N_1N.p_data.csv


Data associated with
├── fig_2b

Ranked gene set enrichment results, ranking genes by signed z-score from per-gene DGE models (week 14 PR vs. week 0 PR, adjusted for cell composition) in ascending order to test for downregulated gene sets.
│   ├── dream.gene_expr_filtered_vobj_fit_dge_5.topTable_C_3R_1R.gene_set_enrichment.tmodCERNOtest_result_LI_ascending.csv
Ranked gene set enrichment results, ranking genes by signed z-score from per-gene DGE models (week 14 PR vs. week 0 PR, adjusted for cell composition) in descending order to test for upregulated gene sets.
│   └── dream.gene_expr_filtered_vobj_fit_dge_5.topTable_C_3R_1R.gene_set_enrichment.tmodCERNOtest_result_LI_descending.csv

Ranked gene set enrichment results, ranking genes by signed z-score from per-gene DGE models (week 14 PNR vs. week 0 PNR, adjusted for cell composition) in ascending order to test for downregulated gene sets.
│   ├── dream.gene_expr_filtered_vobj_fit_dge_5.topTable_C_3N_1N.gene_set_enrichment.tmodCERNOtest_result_LI_ascending.csv
Ranked gene set enrichment results, ranking genes by signed z-score from per-gene DGE models (week 14 PNR vs. week 0 PNR, adjusted for cell composition) in descending order to test for upregulated gene sets.
│   ├── dream.gene_expr_filtered_vobj_fit_dge_5.topTable_C_3N_1N.gene_set_enrichment.tmodCERNOtest_result_LI_descending.csv

Ranked gene set enrichment results, ranking genes by signed z-score from per-gene DGE models (week 14/week 0 x PR/PNR interaction, adjusted for cell composition) in ascending order to test for downregulated gene sets.
│   ├── dream.gene_expr_filtered_vobj_fit_dge_5.topTable_C_(3R_1R)_(3N_1N).gene_set_enrichment.tmodCERNOtest_result_LI_ascending.csv
Ranked gene set enrichment results, ranking genes by signed z-score from per-gene DGE models (week 14/week 0 x PR/PNR interaction, adjusted for cell composition) in descending order to test for upregulated gene sets.
│   ├── dream.gene_expr_filtered_vobj_fit_dge_5.topTable_C_(3R_1R)_(3N_1N).gene_set_enrichment.tmodCERNOtest_result_LI_descending.csv


Data associated with
├── fig_3b

Per-gene summary statistics from DGE model: natural cubic spline of expression as a function of study day, testing the PR/PNR x study day interaction, adjusted for cell composition.
│   ├── dream.gene_expr_filtered_vobj_fit_dge_3.topTable_coefs_22,23,24.csv

Cluster assignments from hierarchical clustering of significant genes from the above spline model by expression.
│   └── plot_gene_set_enrichment.dge_spline_cluster_genes.csv


Data associated with
├── fig_s3a

Per-gene summary statistics from DGE model: week 0 PR IFX vs. week 0 PNR IFX, unadjusted for cell composition.
│   ├── dream.gene_expr_filtered_vobj_fit_dge_2.topTable_C_1RI_1NI.csv
Per-gene summary statistics from DGE model: week 0 PR ADA vs. week 0 PNR ADA, unadjusted for cell composition.
│   ├── dream.gene_expr_filtered_vobj_fit_dge_2.topTable_C_1RA_1NA.csv
Per-gene summary statistics from DGE model: week 0 PR vs. week 0 PNR, unadjusted for cell composition.
│   └── dream.gene_expr_filtered_vobj_fit_dge_6.topTable_C_1R_1N.csv


Data associated with
├── fig_s3b

Ranked gene set enrichment results, ranking genes by signed z-score from per-gene DGE models (week 0 PR IFX vs. week 0 PNR IFX, unadjusted for cell composition) in ascending order to test for downregulated gene sets.
│   ├── dream.gene_expr_filtered_vobj_fit_dge_2.topTable_C_1RI_1NI.gene_set_enrichment.tmodCERNOtest_result_LI_ascending.csv
Ranked gene set enrichment results, ranking genes by signed z-score from per-gene DGE models (week 0 PR IFX vs. week 0 PNR IFX, unadjusted for cell composition) in descending order to test for upregulated gene sets.
│   ├── dream.gene_expr_filtered_vobj_fit_dge_2.topTable_C_1RI_1NI.gene_set_enrichment.tmodCERNOtest_result_LI_descending.csv

Ranked gene set enrichment results, ranking genes by signed z-score from per-gene DGE models (week 0 PR ADA vs. week 0 PNR ADA, unadjusted for cell composition) in ascending order to test for downregulated gene sets.
│   ├── dream.gene_expr_filtered_vobj_fit_dge_2.topTable_C_1RA_1NA.gene_set_enrichment.tmodCERNOtest_result_LI_ascending.csv
Ranked gene set enrichment results, ranking genes by signed z-score from per-gene DGE models (week 0 PR ADA vs. week 0 PNR ADA, unadjusted for cell composition) in descending order to test for upregulated gene sets.
│   ├── dream.gene_expr_filtered_vobj_fit_dge_2.topTable_C_1RA_1NA.gene_set_enrichment.tmodCERNOtest_result_LI_descending.csv

Ranked gene set enrichment results, ranking genes by signed z-score from per-gene DGE models (week 0 PR/PNR x IFX/ADA interaction, unadjusted for cell composition) in ascending order to test for downregulated gene sets.
│   ├── dream.gene_expr_filtered_vobj_fit_dge_2.topTable_C_(1RI_1NI)_(1RA_1NA).gene_set_enrichment.tmodCERNOtest_result_LI_ascending.csv
Ranked gene set enrichment results, ranking genes by signed z-score from per-gene DGE models (week 0 PR/PNR x IFX/ADA interaction, unadjusted for cell composition) in descending order to test for upregulated gene sets.
│   ├── dream.gene_expr_filtered_vobj_fit_dge_2.topTable_C_(1RI_1NI)_(1RA_1NA).gene_set_enrichment.tmodCERNOtest_result_LI_descending.csv

Ranked gene set enrichment results, ranking genes by signed z-score from per-gene DGE models (week 0 PR vs. week 0 PNR, unadjusted for cell composition) in ascending order to test for downregulated gene sets.
│   ├── dream.gene_expr_filtered_vobj_fit_dge_6.topTable_C_1R_1N.gene_set_enrichment.tmodCERNOtest_result_LI_ascending.csv
Ranked gene set enrichment results, ranking genes by signed z-score from per-gene DGE models (week 0 PR vs. week 0 PNR, unadjusted for cell composition) in descending order to test for upregulated gene sets.
│   └── dream.gene_expr_filtered_vobj_fit_dge_6.topTable_C_1R_1N.gene_set_enrichment.tmodCERNOtest_result_LI_descending.csv


Data associated with
├── fig_s6a

Ranked gene set enrichment results, ranking genes by signed z-score from per-gene DGE models (week 14 PR IFX vs. week 0 PR IFX, adjusted for cell composition) in ascending order to test for downregulated gene sets.
│   ├── dream.gene_expr_filtered_vobj_fit_dge_1.topTable_C_3RI_1RI.gene_set_enrichment.tmodCERNOtest_result_LI_ascending.csv
Ranked gene set enrichment results, ranking genes by signed z-score from per-gene DGE models (week 14 PR IFX vs. week 0 PR IFX, adjusted for cell composition) in descending order to test for upregulated gene sets.
│   ├── dream.gene_expr_filtered_vobj_fit_dge_1.topTable_C_3RI_1RI.gene_set_enrichment.tmodCERNOtest_result_LI_descending.csv

Ranked gene set enrichment results, ranking genes by signed z-score from per-gene DGE models (week 14 PR ADA vs. week 0 PR ADA, adjusted for cell composition) in ascending order to test for downregulated gene sets.
│   ├── dream.gene_expr_filtered_vobj_fit_dge_1.topTable_C_3RA_1RA.gene_set_enrichment.tmodCERNOtest_result_LI_ascending.csv
Ranked gene set enrichment results, ranking genes by signed z-score from per-gene DGE models (week 14 PR ADA vs. week 0 PR ADA, adjusted for cell composition) in descending order to test for upregulated gene sets.
│   ├── dream.gene_expr_filtered_vobj_fit_dge_1.topTable_C_3RA_1RA.gene_set_enrichment.tmodCERNOtest_result_LI_descending.csv

Ranked gene set enrichment results, ranking genes by signed z-score from per-gene DGE models (week 14/week 0 x IFX/ADA interaction in PRs, adjusted for cell composition) in ascending order to test for downregulated gene sets.
│   ├── dream.gene_expr_filtered_vobj_fit_dge_1.topTable_C_(3RI_1RI)_(3RA_1RA).gene_set_enrichment.tmodCERNOtest_result_LI_ascending.csv
Ranked gene set enrichment results, ranking genes by signed z-score from per-gene DGE models (week 14/week 0 x IFX/ADA interaction in PRs, adjusted for cell composition) in descending order to test for upregulated gene sets.
│   ├── dream.gene_expr_filtered_vobj_fit_dge_1.topTable_C_(3RI_1RI)_(3RA_1RA).gene_set_enrichment.tmodCERNOtest_result_LI_descending.csv

Ranked gene set enrichment results, ranking genes by signed z-score from per-gene DGE models (week 14 PR vs. week 0 PR, adjusted for cell composition) in ascending order to test for downregulated gene sets.
│   ├── dream.gene_expr_filtered_vobj_fit_dge_5.topTable_C_3R_1R.gene_set_enrichment.tmodCERNOtest_result_LI_ascending.csv
Ranked gene set enrichment results, ranking genes by signed z-score from per-gene DGE models (week 14 PR vs. week 0 PR, adjusted for cell composition) in descending order to test for upregulated gene sets.
│   └── dream.gene_expr_filtered_vobj_fit_dge_5.topTable_C_3R_1R.gene_set_enrichment.tmodCERNOtest_result_LI_descending.csv


Data associated with
└── fig_s6b

Ranked gene set enrichment results, ranking genes by signed z-score from per-gene DGE models (week 14 PNR IFX vs. week 0 PNR IFX, adjusted for cell composition) in ascending order to test for downregulated gene sets.
    ├── dream.gene_expr_filtered_vobj_fit_dge_1.topTable_C_3NI_1NI.gene_set_enrichment.tmodCERNOtest_result_LI_ascending.csv
Ranked gene set enrichment results, ranking genes by signed z-score from per-gene DGE models (week 14 PNR IFX vs. week 0 PNR IFX, adjusted for cell composition) in descending order to test for upregulated gene sets.
    ├── dream.gene_expr_filtered_vobj_fit_dge_1.topTable_C_3NI_1NI.gene_set_enrichment.tmodCERNOtest_result_LI_descending.csv

Ranked gene set enrichment results, ranking genes by signed z-score from per-gene DGE models (week 14 PNR ADA vs. week 0 PNR ADA, adjusted for cell composition) in ascending order to test for downregulated gene sets.
    ├── dream.gene_expr_filtered_vobj_fit_dge_1.topTable_C_3NA_1NA.gene_set_enrichment.tmodCERNOtest_result_LI_ascending.csv
Ranked gene set enrichment results, ranking genes by signed z-score from per-gene DGE models (week 14 PNR ADA vs. week 0 PNR ADA, adjusted for cell composition) in descending order to test for upregulated gene sets.
    ├── dream.gene_expr_filtered_vobj_fit_dge_1.topTable_C_3NA_1NA.gene_set_enrichment.tmodCERNOtest_result_LI_descending.csv

Ranked gene set enrichment results, ranking genes by signed z-score from per-gene DGE models (week 14/week 0 x IFX/ADA interaction in PNRs, adjusted for cell composition) in ascending order to test for downregulated gene sets.
    ├── dream.gene_expr_filtered_vobj_fit_dge_1.topTable_C_(3NI_1NI)_(3NA_1NA).gene_set_enrichment.tmodCERNOtest_result_LI_ascending.csv
Ranked gene set enrichment results, ranking genes by signed z-score from per-gene DGE models (week 14/week 0 x IFX/ADA interaction in PNRs, adjusted for cell composition) in descending order to test for upregulated gene sets.
    ├── dream.gene_expr_filtered_vobj_fit_dge_1.topTable_C_(3NI_1NI)_(3NA_1NA).gene_set_enrichment.tmodCERNOtest_result_LI_descending.csv

Ranked gene set enrichment results, ranking genes by signed z-score from per-gene DGE models (week 14 PNR vs. week 0 PNR, adjusted for cell composition) in ascending order to test for downregulated gene sets.
    ├── dream.gene_expr_filtered_vobj_fit_dge_5.topTable_C_3N_1N.gene_set_enrichment.tmodCERNOtest_result_LI_ascending.csv
Ranked gene set enrichment results, ranking genes by signed z-score from per-gene DGE models (week 14 PNR vs. week 0 PNR, adjusted for cell composition) in descending order to test for upregulated gene sets.
    └── dream.gene_expr_filtered_vobj_fit_dge_5.topTable_C_3N_1N.gene_set_enrichment.tmodCERNOtest_result_LI_descending.csv
